# Multi-Omics Modeling Reveals Peripheral Signatures of Non-Suicidal Self-Injury in Adolescents

**DOI:** 10.64898/2026.07.20.26358487

**Authors:** Fangyi Zhao, Yihang Bao, Wenjing Liu, Tianqi Liu, Weidi Wang, Zhen Liu, Xiaoxia Lei, Xifeng Xia, Wenhong Cheng, Guan Ning Lin

## Abstract

Non-suicidal self-injury (NSSI) is common among adolescents with emotional disorders, yet biological indicators of current NSSI status remain limited. We developed a genome-aware multi-omics modeling framework in 107 adolescents with emotional disorders, including 53 without NSSI and 54 with current NSSI. The model integrated metabolomic, inflammatory, clinical blood and genome-derived features, with polygenic risk score and rare variant burden used as genetic-context variables. The fusion model achieved the strongest classification performance (mean AUC = 0.811) and outperformed single-omics alternatives, indicating that NSSI status was better represented by distributed multi-omics patterns than by a single biomarker layer. Repeated modeling prioritized 42 stable features, many of which were not significant in conventional univariate testing. Group-specific network reconstruction further revealed peripheral reorganization, including convergence of non-NSSI modules into an NSSI-associated module that linked inflammatory recruitment with weaker immune-communication, repair and support-related signals. Exploratory MRI, gut-related and stress-endocrine analyses provided additional biological anchors, while a compact sentinel marker panel translated the full model into clinically readable profiles. These findings support a distributed, genome-aware peripheral state associated with current NSSI and provide a framework for future validation of multi-omics state markers in adolescent emotional disorders.

**Biographical Note:** Guan Ning Lin is Professor at the School of Biomedical Engineering, Shanghai Jiao Tong University; Vice Director of Imaging, Computing and Systems Biomedicine; Director of Shanghai Gener Arfysica Intelligent Healthcare & Brain Science Research Institute; Deputy Director of MOE Digital Medicine Engineering Center; and Researcher at Shanghai Mental Health Center.

## 1. Introduction

Non-suicidal self-injury (NSSI) is a major clinical concern in adolescents with emotional disorders [1, 2]. It is closely associated with difficulties in affect regulation and broader symptom burden [3, 4], frequently co-occurs with adolescent depression requiring targeted intervention [5], and longitudinal evidence links self-injurious thoughts and behaviors to later suicidal ideation, suicide attempts and suicide death [6]. In clinical practice, current NSSI status is still identified mainly through interviews, behavioral history and self-report [7]. These approaches are essential, but they provide only indirect access to the biological condition that accompanies current NSSI. Objective and interpretable biological indicators may therefore complement clinical assessment, particularly when the aim is to identify current state rather than lifetime history [8].

Prior work has implicated inflammation in NSSI and related adolescent psychopathology [9, 10], together with stress-endocrine physiology [11, 12], pain-related neurophysiology [13, 14], and neuroinflammatory or synaptic-support biology [15, 16]. Metabolic and gut-related pathways may also be relevant in adolescent depression and NSSI-related contexts [17, 18]. Findings from single molecules or single omics layers, however, have remained inconsistent [19, 13]. One explanation is that current NSSI status is not dominated by one biomarker, but is carried by several weak-to-moderate signals that become informative only when considered together. This makes a multi-omics design better suited than a single-marker approach for capturing coordinated biological variation [20, 21].

Genetic background may also change how peripheral molecular signals should be read. The same inflammatory, metabolic or blood-based profile may imply different NSSI-state probabilities depending on inherited liability, rare variant burden or interactions between genomic context and omics layers. This point is relevant for NSSI and self-harm phenotypes, where common genetic architecture has been linked to child psychopathology and brain structure [22], and broader suicide-attempt liability overlaps with psychiatric and behavioral risk dimensions [23, 24]. Cross-population PRS methods and multi-trait summary-statistic approaches now make it possible to model this genetic context more explicitly [25, 26]. A genome-aware multi-omics model can therefore test whether peripheral omics information gains meaning when interpreted together with PRS, rare burden and genetic-by-omics interaction terms.

Classification performance alone is not enough for biological interpretation. A model may distinguish current NSSI status, but an AUC value does not show whether the informative features form an organized biological structure. Precision-psychiatry work therefore emphasizes interpretable modeling, not prediction alone [20, 21]. Recent machine-learning studies in NSSI have reached a similar conclusion, highlighting the need for validation and interpretability [27, 28]. To move beyond black-box classification, stable model-prioritized features can be examined through group-specific network reconstruction. This analysis asks whether repeatedly selected features are coordinated differently in NSSI and non-NSSI states, whether module membership is reorganized and whether particular network regions carry interpretable peripheral biology [29, 30].

Here, we developed a genome-aware multi-omics stacking model to identify current NSSI status in adolescents with emotional disorders. The model integrated metabolomic, inflammatory, clinical blood and genome-derived features, with PRS and rare burden modeled as genetic-context variables. We then prioritized stable model features, reconstructed group-specific feature networks and examined whether NSSI was associated with peripheral network reorganization. White-matter-normalized MRI readouts, gut-related and stress-endocrine features, and a compact sentinel marker panel were used for exploratory anchoring and clinical-readability analyses.

## 2. Results

### 2.1 Multi-omics fusion identifies a distributed signature of current NSSI status

We first tested whether current NSSI status could be identified from a distributed multi-omics signature rather than from a single molecular layer (Fig. 1). The final multi-omics cohort included 107 adolescents with emotional disorders, including 53 non-NSSI participants and 54 participants with NSSI. Genome-derived features, metabolomic features, inflammatory factors and clinical blood indicators were used for model construction, while scale-derived phenotypes and MRI readouts were reserved for downstream exploratory biological anchoring. Modality-specific expert models were combined through a stacking logistic regression model to integrate complementary information across molecular systems (Fig. 2a).

**Figure 1.**
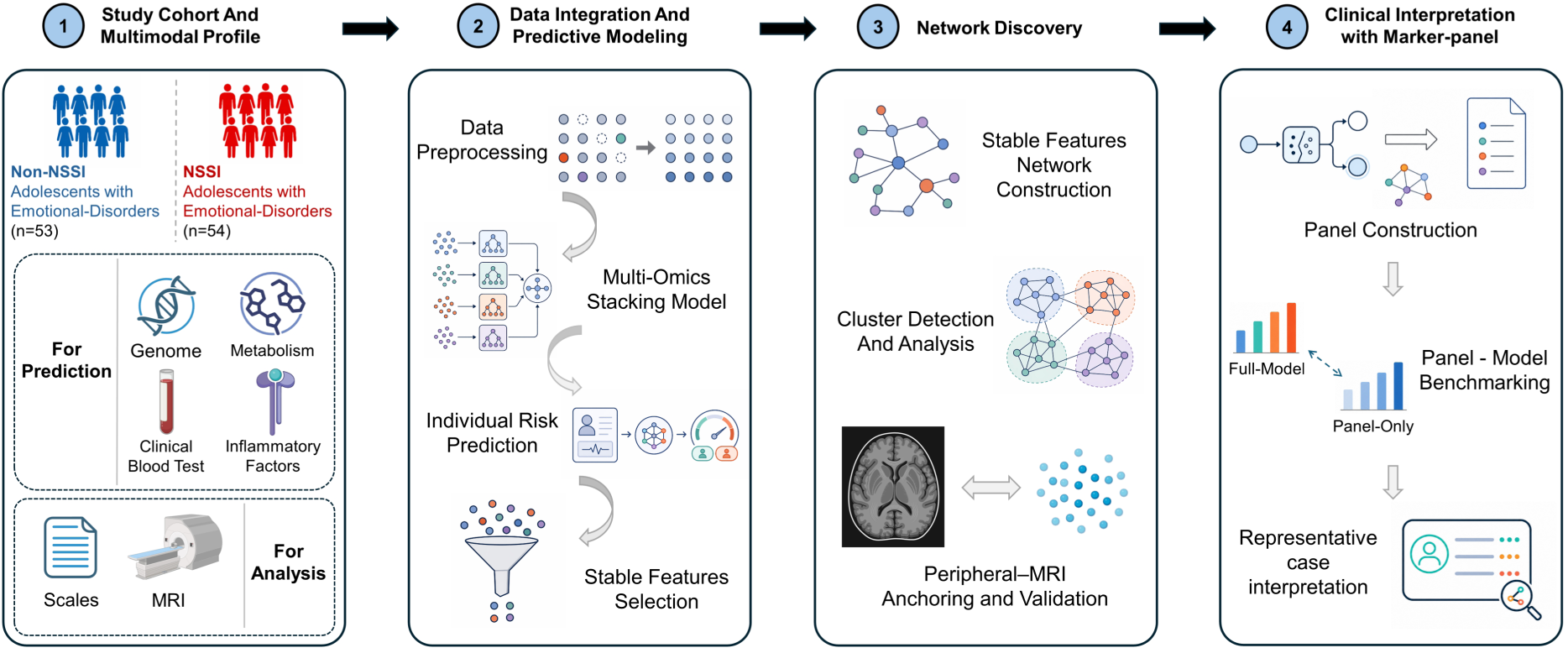
Study design and analytical workflow. Overview of cohort construction, genome-aware multi-omics modeling, stable-feature prioritization, network analysis, exploratory biological anchoring and sentinel-panel interpretation. NSSI, non-suicidal self-injury; MRI, magnetic resonance imaging.

**Figure 2.**
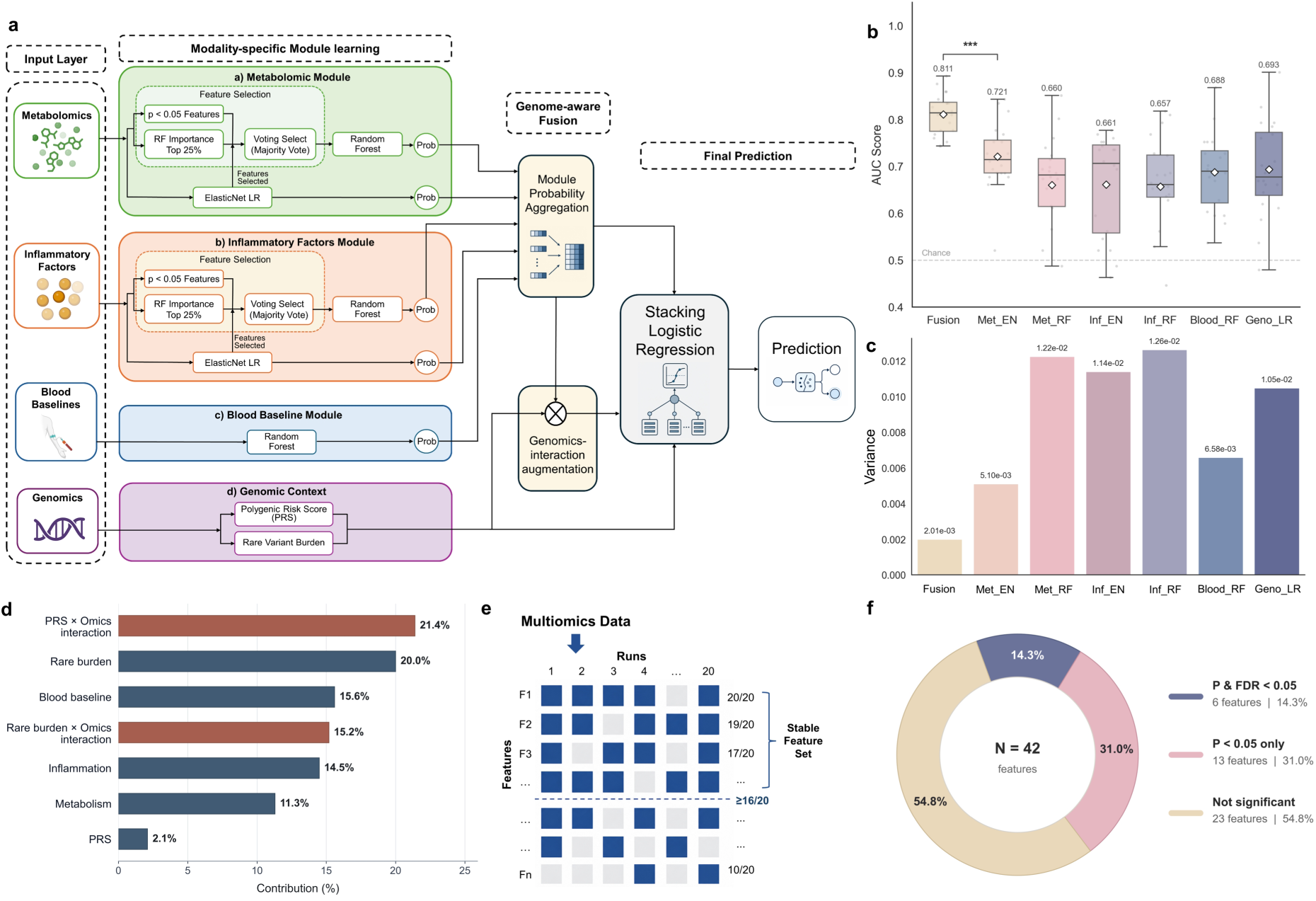
Multi-omics model performance. a, Multi-omics stacking framework integrating metabolomic, inflammatory, blood and genome-derived information. b,c, AUC performance and variance across repeated splits. d, Meta-model contribution summary. e,f, Stable-feature selection and univariate significance profile.

The integrated fusion model showed the strongest classification performance among the evaluated models (Fig. 2b,c). It reached an AUC of 0.811, whereas individual expert models showed lower AUC values, including metabolomic ElasticNet (0.721), metabolomic random forest (0.660), inflammatory ElasticNet (0.661), inflammatory random forest (0.657), blood random forest (0.688), and genomic logistic regression (0.693). The AUC variance was also lowest for the fusion model (2.01e-03), compared with higher variance values for the single-omics models. This pattern suggests that current NSSI status was not best represented by one dominant molecular source. Instead, discrimination relied on weak-to-moderate signals distributed across metabolic, inflammatory, clinical blood and genetic-context dimensions, consistent with biology-informed multi-omics modeling approaches [20, 21].

We next used modality-level ablation analyses to test whether fusion performance depended on specific omics components, with numerical results summarized in Table 1. Removing metabolomic features reduced the mean AUC from 0.811 to 0.755 (P = 0.0093), and removing blood baseline features reduced it to 0.781 (P = 0.0037). Removing genomic context produced a further drop to 0.761 (P < 0.001), consistent with a substantial contribution from genome-aware integration. Removing inflammatory factors produced only a modest AUC change (0.802, P = 0.1851). Although Brier score differences were statistically significant across ablation settings, the absolute calibration changes were small for some modalities, particularly inflammatory factors. The ablation results therefore mainly support distributed discrimination, with calibration effects interpreted as supportive rather than definitive.

**Table 1.**
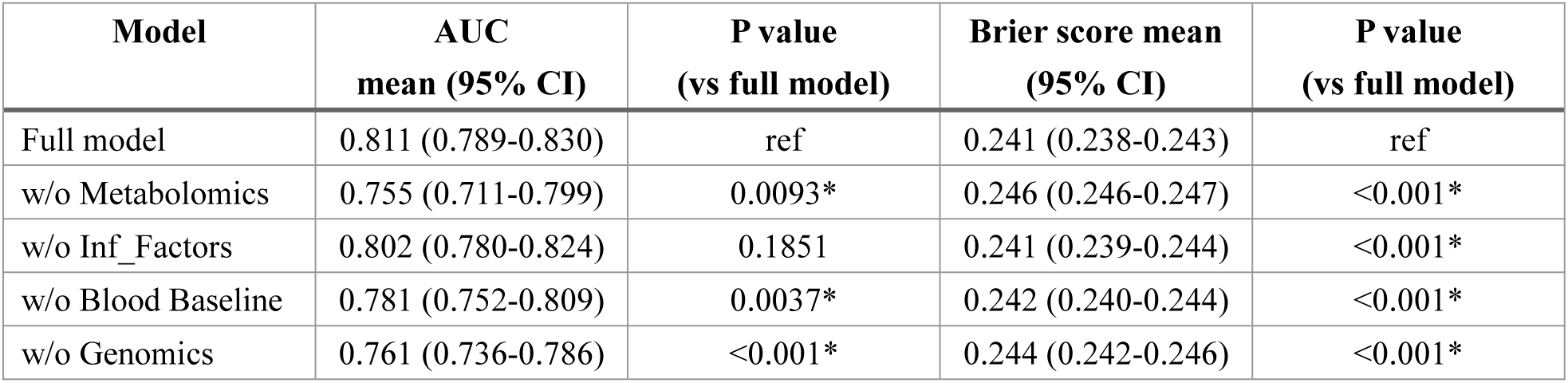
Omics ablation results.

The coefficient-based meta-model contribution summary indicated that both genetic context and non-genetic omics layers contributed to classification (Fig. 2d). PRS-by-omics interaction terms contributed 21.4%, rare burden contributed 20.0%, blood baseline features contributed 15.6%, rare burden-by-omics interaction terms contributed 15.2%, inflammatory factors contributed 14.5%, metabolomic features contributed 11.3%, and PRS alone contributed 2.1%. Two points are worth noting. First, genetic-context features were informative as stand-alone predictors and through interaction terms with omics expert outputs, consistent with genome-aware psychiatric modeling concepts [20, 21]. Second, inflammatory and metabolic features remained informative in the stacked model even when their isolated expert performance was modest. The model therefore captured a genome-aware peripheral profile in which the same non-genetic molecular pattern may carry different NSSI-related meaning depending on genetic background, a possibility supported by recent NSSI and suicide-genetics studies [22, 24].

Repeated modeling prioritized 42 stable features for downstream interpretation (Fig. 2e,f). These features were not equivalent to a conventional univariate biomarker list: 6 of 42 features (14.3%) met both nominal P and FDR criteria, 13 features (31.0%) reached nominal P < 0.05 without surviving FDR correction, and 23 features (54.8%) were not significant in the univariate summary. The model therefore retained many features that were not individually significant but were repeatedly informative in the multivariate setting. We reasoned that their biological relevance might lie in coordinated, context-dependent relationships among molecular systems, rather than in large independent mean shifts. This motivated a group-specific network analysis to ask whether the model had captured an organized peripheral biological pattern.

### 2.2 Stable-feature networks reveal peripheral reorganization

Using the 42 stable features, with PRS_score and Rare_Burden displayed as genetic-context nodes in Fig. 3a, we constructed group-specific feature networks for NSSI and non-NSSI participants to determine whether model-prioritized features were organized differently across groups. The two networks showed comparable overall feature representation but distinct architecture (Fig. 3a). This argues against a simple interpretation in which the NSSI network reflects a nonspecific global collapse of molecular coordination. Instead, NSSI-related differences appeared as network rewiring: feature associations were reassigned, module membership changed, and the features occupying bridge-like positions differed between groups.

**Figure 3.**
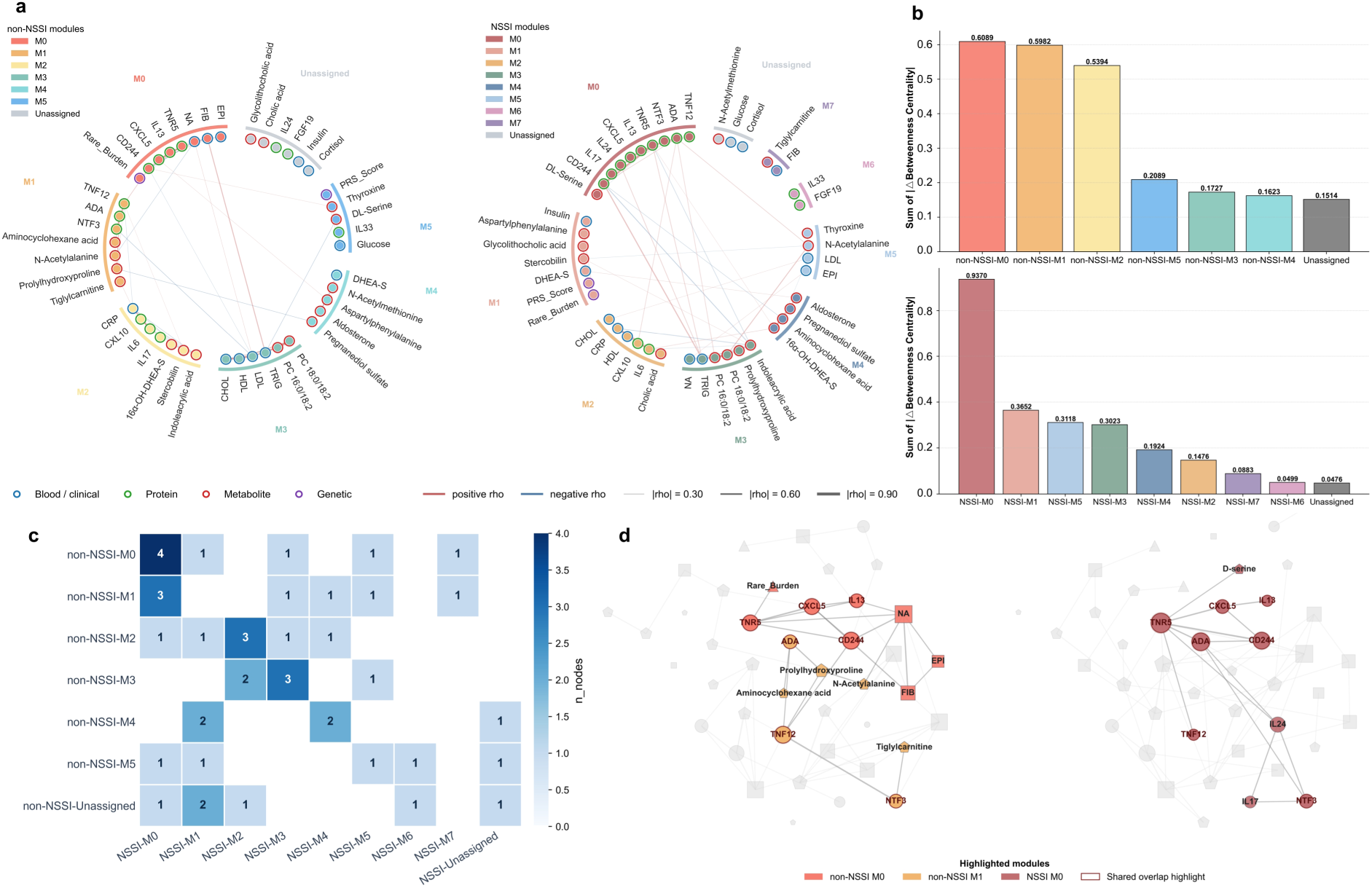
Peripheral network reorganization. a, Group-specific stable-feature networks in non-NSSI and NSSI participants, with PRS_score and Rare_Burden displayed as genetic-context nodes. b, Module-level centrality reconfiguration. c, Module-transition heatmap. d, Matched network view showing reassignment of non-NSSI M0/M1 features into NSSI-M0.

To identify the network regions most affected by this reorganization, we quantified module-level centrality changes based on betweenness centrality (Fig. 3b). Modules with larger changes were prioritized because altered betweenness suggests that a feature, or a set of features, has changed its role in connecting otherwise separated parts of the network. This analysis highlighted NSSI-M0 as a prominently reconfigured module, while non-NSSI M0 and non-NSSI M1 also showed marked centrality changes. We therefore treated these modules as candidate regions of biological reorganization, not as isolated biomarker clusters.

We then examined the module transition heatmap together with the matched network view to determine how these reconfigured modules were related across states (Fig. 3c,d). The heatmap showed that features assigned to non-NSSI M0 and M1 were substantially reassigned into NSSI-M0. The matched network view made this relationship visible: features distributed across two non-NSSI modules became concentrated within one NSSI-associated module. This transition was biologically informative because it suggested that separable non-NSSI feature environments became topologically compressed into a single NSSI-M0-centered structure. NSSI-M0 was therefore prioritized not simply because it contained several differential features, but because it represented a module-reassignment event involving non-NSSI M0 and M1 together with a prominent shift in centrality structure.

These structural analyses showed that current NSSI status was associated with reorganization of the stable-feature network. The model-prioritized features formed an interpretable peripheral pattern, not a disconnected list of markers. We next examined whether the reorganized NSSI-M0 structure supported a more specific biological interpretation.

### 2.3 A reconfigured peripheral module links inflammatory activation to support and repair insufficiency

Within the reorganized NSSI-associated subnetwork, ADA occupied a bridge-like position that linked module-transition structure to immune-communication and repair-related features (Fig. 4a). ADA was positioned close to CD244 and TNFRSF5/CD40, placing purinergic-related variation within a broader immune-communication context rather than as an isolated metabolic signal. Because of space constraints, TNFRSF5/CD40 is abbreviated as TNR5 in the figures, and TNFSF12/TWEAK is abbreviated as TNF12. The NSSI-M0-centered module contained ADA, CD244, CXCL5, D-serine, IL13, IL17, IL24, NTF3, TNFSF12/TWEAK and TNFRSF5/CD40. Rather than representing a single inflammatory marker or a generic inflammatory cluster, this module connected inflammatory recruitment, immune communication, and support and remodeling-related signals, represented mainly by CXCL5/IL24, ADA/CD244/TNFRSF5 and NTF3/D-serine/TNFSF12, respectively.

**Figure 4.**
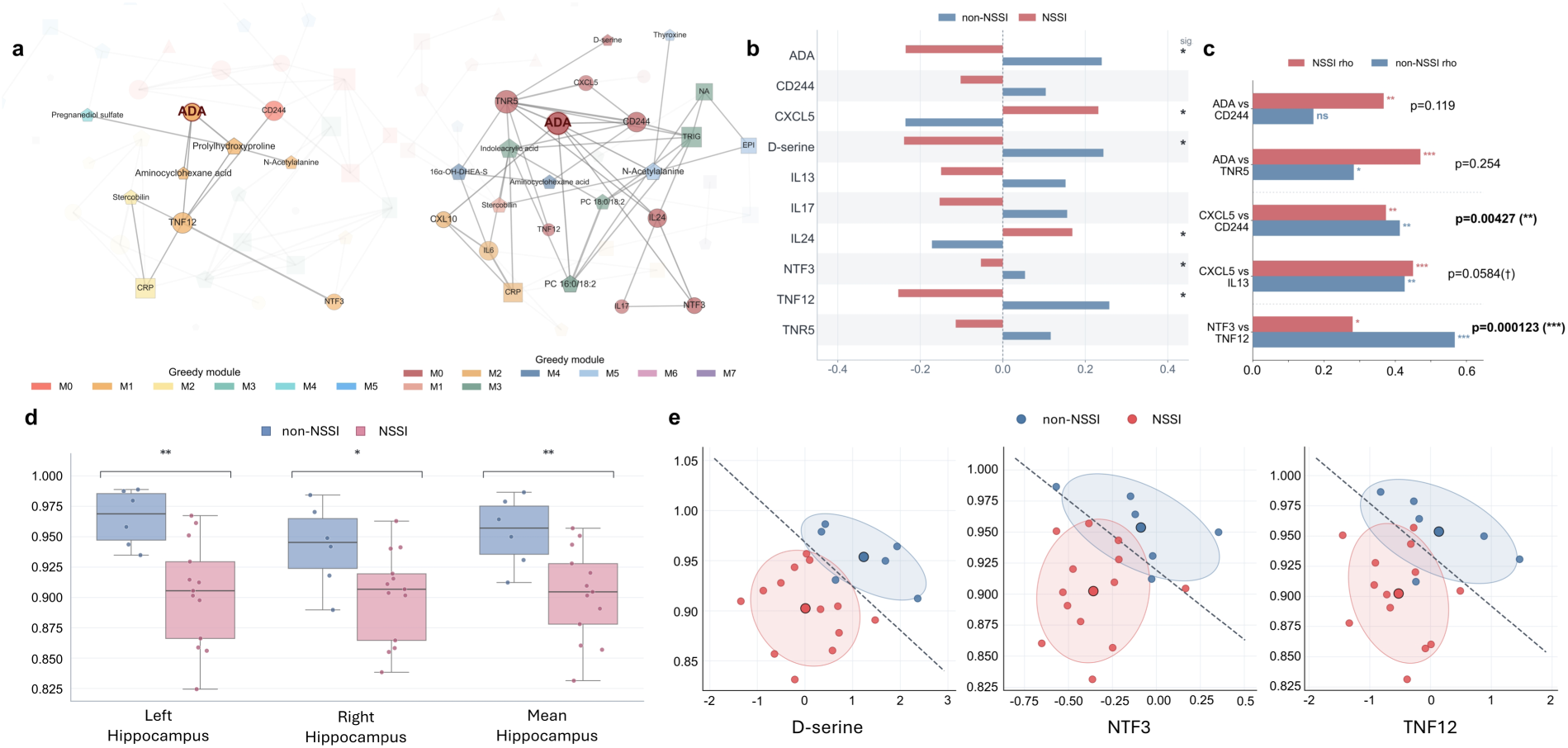
Inflammatory-support module. a, Reconfigured NSSI-associated subnetwork. b, Group-mean directions of recruitment, immune-communication and support/remodeling markers. c, Selected group-specific feature associations. d,e, Exploratory MRI anchoring of support-related markers using white-matter-normalized hippocampal readouts.

Feature-level directions supported this interpretation (Fig. 4b). CXCL5 and IL24 were shifted upward in NSSI, whereas ADA, CD244, D-serine, IL13, IL17, NTF3, TNFSF12/TWEAK and TNFRSF5/CD40 were shifted downward or showed lower group means in NSSI. This was not a simple “inflammation high” profile, even though inflammatory markers have been associated with adolescent NSSI [9, 10]. A more precise description is inflammatory activation accompanied by weaker support and repair capacity: recruitment-type signals were relatively prominent, while several immune-communication, remodeling, neurotrophic and synaptic-support features were reduced. In this interpretation, the biologically relevant signal is the coexistence of inflammatory recruitment with weaker support and repair capacity in the same NSSI-M0-centered pattern, rather than the absolute direction of any single marker. This makes NSSI-M0 a peripheral-state model, not a conventional single-marker inflammatory signature, consistent with cautions raised by biomarker studies in mental health [19].

Pairwise association analyses further supported this interpretation (Fig. 4c). ADA remained positively related to CD244 and TNFRSF5/CD40 in the NSSI group, but interaction tests for these ADA-centered relationships were not robust (P = 0.119 and P = 0.254, respectively). By contrast, the CXCL5-CD244 relationship showed a significant group-by-feature interaction (interaction P = 0.00427), and the CXCL5-IL13 relationship showed a weaker trend-level interaction (interaction P = 0.0584). These results suggest that the relationship between inflammatory recruitment and immune-communication signals was reorganized in NSSI, not merely amplified. The strongest pairwise evidence involved the NTF3-by-group interaction for TNFSF12/TWEAK (interaction P = 0.000123). Because NTF3 represents neurotrophic support and D-serine represents NMDA receptor-dependent synaptic support, whereas TNFSF12/TWEAK is related to tissue remodeling and immune-neurovascular interface biology, the altered NTF3-TNFSF12/TWEAK relationship indicates weakened support and repair coupling within the NSSI-M0-centered module.

This organization supports an exploratory peripheral-limbic working model. In adolescents with emotional disorders, repeated stress exposure may bias endocrine background [11, 12], which can coexist with stronger inflammatory recruitment and altered neuroimmune signaling [15, 16]. Weaker purinergic and NK or T-cell immune communication, together with altered tissue-remodeling signals, may influence blood-brain barrier, neurovascular and neuroimmune interfaces. Lower NTF3 and D-serine also point to weaker neurotrophic and NMDA-dependent synaptic-support biology. Brain and pain-physiology studies of NSSI support cautious cross-system interpretation [14, 31]. In this exploratory model, the peripheral module and limbic readouts are interpreted as potentially connected components of a stress-sensitive biological state, not as a linear one-way pathway [32, 33].

In an exploratory MRI subset analysis, we asked whether the support component of this peripheral signature showed concordance with brain-end readouts. After MRI quality control, sample matching and outlier removal, white-matter-normalized hippocampal signal ratios were lower in the NSSI group across left, right and mean hippocampal measures (Fig. 4d). These dimensionless ratios were calculated as hippocampal ROI mean intensity divided by the participant-specific white-matter mean intensity. The MRI readouts were used as exploratory peripheral-limbic anchors for the NTF3/D-serine support component, not as mechanistic imaging biomarkers.

The two-dimensional MRI anchoring plots showed partial separation and directional alignment between support-related markers and hippocampus-to-WM ratios (Fig. 4e). The same peripheral module that suggested inflammatory activation with weaker support and repair capacity also contained support-related features that aligned with white-matter-normalized hippocampal readouts. This cross-scale concordance should be interpreted cautiously because of the small MRI subset, but it provides exploratory support for linking the peripheral support arm of the network to limbic-system variation relevant to stress contextualization, affective memory and behavioral regulation [32, 33].

### 2.4 Exploratory gut-related and stress-endocrine features define physiological context

We also examined gut-related and stress-endocrine features as exploratory physiological context for the reconfigured peripheral module (Fig. 5). These analyses were intended to place the inflammatory-support pattern in a broader biological setting, not to introduce a separate competing mechanism.

**Figure 5.**
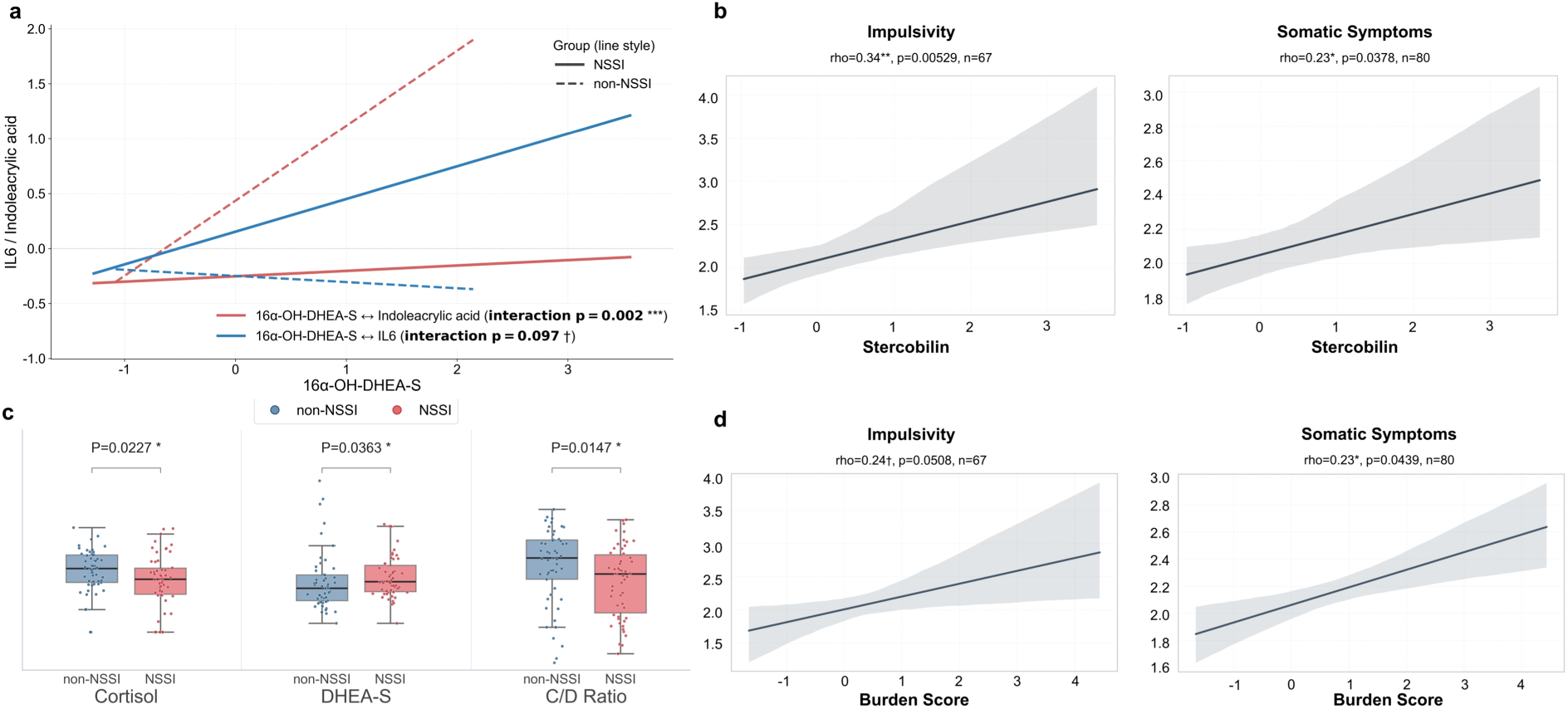
Gut-endocrine physiological context. a, Group-specific coupling among steroid-related, gut-derived and inflammatory features. b, Associations between stercobilin and symptom dimensions. c, Stress-endocrine group comparisons. d, Associations between hormone burden and symptom dimensions.

The gut-related panel showed group-specific coupling among steroid-related metabolites, gut-derived metabolites and inflammatory signals. The interaction between 16alpha-OH-DHEA-S and trans-3-indoleacrylic acid differed between groups (interaction P = 0.002), whereas the interaction involving IL6 showed a weaker trend (interaction P = 0.097) (Fig. 5a). This pattern suggests that gut-derived indole metabolism and steroid-related stress biology may be coordinated differently in NSSI. Physiologically, these features may influence the main NSSI-M0 pattern through two plausible routes: gut-derived metabolites can modulate immune tone [34, 18], while steroid-related signals can alter immune-cell sensitivity and stress-response thresholds [11, 12].

In exploratory symptom-association analyses, stercobilin was positively associated with impulsivity (rho = 0.34, P = 0.00529, n = 67) and somatic symptoms (rho = 0.23, P = 0.0378, n = 80) (Fig. 5b). These associations suggest that gut-related metabolic variation may map onto behavioral and somatic dimensions, instead of serving only as a peripheral metabolic readout.

Stress-endocrine markers also showed exploratory group differences and symptom associations. Cortisol differed between groups (P = 0.0227), DHEA-S differed between groups (P = 0.0363), and the cortisol/DHEA-S (C/D) ratio differed between groups (P = 0.0147) (Fig. 5c). A direction-aligned hormone burden score showed a weak positive correlation with impulsivity that did not reach the nominal 0.05 threshold (rho = 0.24, P = 0.0508, n = 67) and was positively associated with somatic symptoms (rho = 0.23, P = 0.0439, n = 80) (Fig. 5d). These findings suggest that HPA-related and adrenal-steroid variation may provide a stress-endocrine background that biases the peripheral network toward or away from the NSSI-associated pattern [11, 12].

The gut-related and stress-endocrine findings outline a possible physiological background for the reconfigured peripheral module. Gut-derived indole and stercobilin signals, together with C/D ratio and hormone-burden variation, may shape immune communication, stress responsiveness and symptom expression. These contextual features may help explain why a peripheral network pattern could be clinically heterogeneous: similar inflammatory or immune signals may carry different meaning depending on gut-metabolic and endocrine background.

### 2.5 A compact marker panel provides clinically readable profiles

We then asked whether the full multi-omics classifier could be summarized as a clinically readable individual profile. To do this, we constructed a compact sentinel marker panel from model-prioritized and network-anchored features (Fig. 6a). The panel was designed as an exploratory interpretation layer, not as a replacement for the full model, reflecting the need for interpretable biology-informed modeling [20, 21]. Markers were selected to represent four biological dimensions emerging from the reconfigured-module interpretation: inflammatory recruitment, immune communication, support and remodeling-related biology, and stress-endocrine background. The resulting panel included CXCL5 and IL24 for recruitment, ADA for immune communication, TNFSF12/TWEAK, NTF3 and D-serine for support and remodeling, and the cortisol/DHEA-S (C/D) ratio for endocrine context.

**Figure 6.**
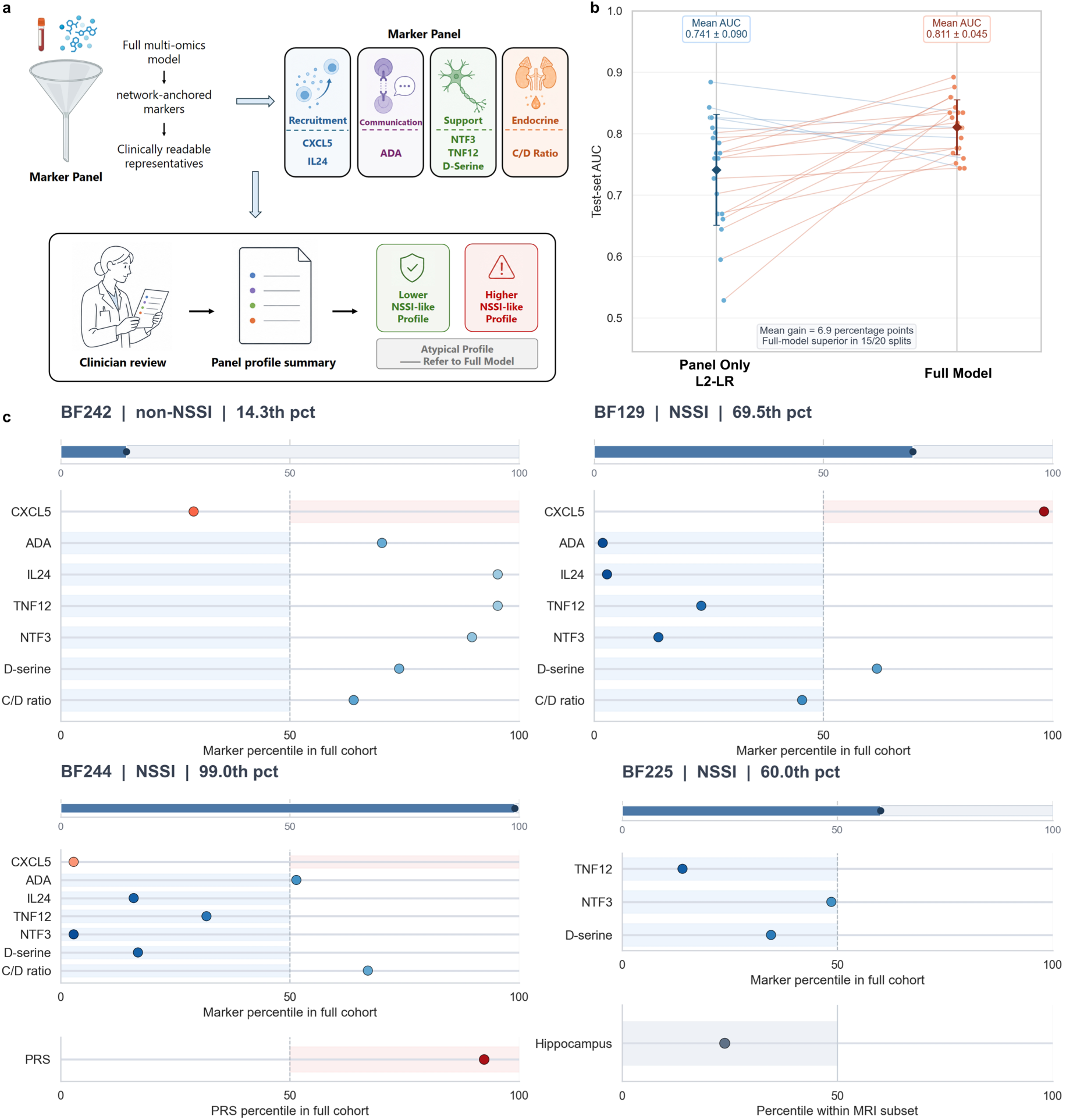
Sentinel marker profiles. a-d, Sentinel-panel construction, benchmarking, representative profiles and probability landscape.

We next benchmarked how much NSSI-state information was retained by this compact panel relative to the full classifier. Using the same repeated train-test splits and an L2-regularized logistic-regression setting matched to the meta-classifier, a panel-only model achieved a mean AUC of 0.741, indicating that the sentinel markers retained part of the model-derived state information (Fig. 6b). The full model nevertheless achieved a higher mean AUC of 0.811 and outperformed the panel-only model in 15 of 20 repeated splits, with a mean gain of 6.9 percentage points. The sentinel panel should therefore be viewed as a readable approximation of the NSSI-associated profile, while the full model retained additional discriminative value from distributed multi-omics and genetic-context information.

Representative cases illustrated how the panel can be interpreted at the individual level (Fig. 6c). BF242, a non-NSSI participant with a low full-model percentile (14.3rd percentile), showed relatively preserved ADA-related communication and support and remodeling-related signals, consistent with a lower NSSI-like profile. In contrast, BF129, an NSSI participant with a higher full-model percentile (69.5th percentile), showed a more NSSI-like configuration, with a strong recruitment signal, reduced ADA-related immune communication, and a partially altered support-related profile. These two cases provided reference profiles at the lower and higher ends of the model-derived NSSI-state spectrum.

The remaining cases highlighted the added value and biological anchoring of the full model. BF244 was an NSSI participant with an extremely high full-model percentile (99.0th percentile) but a less canonical sentinel-panel profile. In the final stacking logistic-regression model, this individual showed a large PRS-by-omics contribution, supporting the interpretation that genetic-context interactions helped the full model classify this panel-discordant profile. BF225 provided an MRI-anchored example: this NSSI participant had an intermediate full-model percentile (60.0th percentile), and reduced support-related markers were accompanied by a lower hippocampus-to-WM ratio percentile within the MRI subset. These cases show that the sentinel panel can make model output clinically readable, while the full model remains necessary for integrating atypical or genetically contextualized profiles. The sentinel panel should therefore be viewed as a clinician-facing summary of the full-model state profile, not as a reduced diagnostic substitute.

## 3. Discussion

This study develops a genome-aware multi-omics model for current NSSI status in adolescents with emotional disorders. The main implication is not that NSSI can be reduced to a single peripheral marker. Rather, current NSSI status may be better understood as a distributed biological condition in which genetic context, immune communication, metabolic background, endocrine stress biology and clinical blood indicators jointly shape model-readable risk. This view fits the clinical heterogeneity of NSSI, which spans developmental trajectories, assessment challenges and psychosocial risk contexts [1, 7]. The same behavior may arise through partially overlapping biological routes, making single-marker designs unlikely to capture the full state space and supporting biology-informed multi-omics approaches [20, 21].

A useful interpretation is to distinguish biological signal from biological cause. The model-prioritized features and group-specific networks identify coordinated peripheral patterns associated with current NSSI status, but they do not establish a causal pathway from inflammation or metabolism to self-injury. Their value is closer to a map of relationships. In that map, inflammatory recruitment, immune-cell communication, and support and remodeling features appear to be coordinated differently in adolescents with current NSSI. This relational view may help explain why biomarker studies of self-harm, depression and adolescent psychopathology have produced mixed single-marker findings: some support inflammatory involvement [9, 10], while broader biomarker work emphasizes heterogeneity and interpretive limits [19]. The informative unit may be the configuration among systems, rather than the isolated level of one molecule.

The peripheral pattern is also relevant to stress-sensitive developmental biology. Adolescence is a period in which stress exposure, affect regulation, endocrine reactivity and immune maturation remain dynamically coupled [11, 12]. A pattern marked by stronger recruitment signals together with weaker support or repair-related signals could plausibly reflect a system that is more reactive than restorative. This pattern may not be specific to NSSI alone, but it may identify a biological context in which emotional distress, impulsivity and somatic burden are more likely to translate into self-injurious behavior, consistent with recent work on maltreatment, impulsivity and clinical assessment [35, 4]. Future work should test whether similar network patterns distinguish NSSI from depression severity, anxiety burden, suicidal ideation or other forms of behavioral dysregulation.

The exploratory MRI and gut-endocrine analyses should be read as hypothesis-generating anchors, not confirmatory mechanisms. Hippocampal readouts are relevant because the hippocampus is sensitive to stress hormones, inflammatory signaling and developmental adversity, but the small MRI subset cannot support strong neurobiological claims. Recent NSSI neuroimaging studies provide a useful context for this cautious interpretation [32, 33]. Similarly, the gut-related and stress-endocrine associations suggest plausible physiological settings in which immune and metabolic signals may vary, but they do not prove a gut-brain or endocrine causal pathway. Gut-brain and microbiota findings in adolescent depression support this hypothesis-generating direction [34, 18]. In the present study, these analyses broaden the interpretation of the multi-omics state and identify cross-system hypotheses for larger, longitudinal and experimentally informed cohorts.

Methodologically, the study illustrates why multi-omics psychiatric modeling needs both predictive and interpretive layers [20, 21]. The stacking model improved classification by allowing genetic context to modify the meaning of non-genetic omics signals, while stable-feature selection and network reconstruction made the classifier more biologically readable. The sentinel marker panel adds a translational layer by giving clinicians and researchers a compact profile with recognizable biological dimensions. Even so, the panel should not be viewed as a diagnostic substitute. Its lower performance relative to the full model is informative because it shows that readability and completeness are not the same. Simplified panels may help communication, but distributed models may still be needed for heterogeneous or genetically contextualized cases, especially given current limitations in NSSI machine-learning prediction [27, 28].

Several limitations should be emphasized. First, the cohort was modest in size and came from a single clinical setting, so external validation is essential before the model or marker panel can be considered generalizable. Second, the study is cross-sectional, which prevents separation of pre-existing vulnerability, current-state correlates and consequences of repeated NSSI or clinical stress. Third, medication exposure, illness duration, pubertal stage, sleep, diet, menstrual cycle and recent stress may influence inflammatory, endocrine and metabolic readouts and should be modeled more deeply in future datasets, particularly given known limitations in inflammatory and HPA-axis biomarker interpretation [19, 12]. Fourth, the MRI subset was small, and the white-matter-normalized signal ratios are relative imaging features rather than quantitative MRI biomarkers. Fifth, network modules depend on feature selection, correlation thresholds and preprocessing choices. They are useful for interpretation, but they should not be treated as fixed biological entities without replication.

Future studies should prioritize independent replication, longitudinal sampling and comparison groups that separate NSSI from broader emotional-disorder severity. Repeated sampling before and after symptom change or treatment would be especially informative, because a clinically useful state marker should vary with NSSI fluctuation instead of merely distinguishing historical groups [8]. Larger imaging datasets could test whether peripheral support and remodeling markers align with stress-sensitive limbic, salience or regulatory circuits [32, 33]. Targeted mechanistic studies will also be needed to determine whether the peripheral relationships highlighted here are causal contributors, compensatory responses or downstream traces of stress and injury-related physiology.

Overall, this study positions adolescent NSSI as a clinically defined behavior that may be accompanied by a measurable, distributed and genetically contextualized biological profile. Its main contribution is to connect classification, network-level interpretation, exploratory cross-system anchoring and clinically readable profile construction in a single analytic workflow. This workflow is not yet a clinical test, but it provides a structured path for moving NSSI biology beyond isolated biomarker searches toward validated state modeling [20, 21].

## 4. Methods

### 4.1 Study participants and clinical assessment

Adolescents were recruited from or assessed at Shanghai Mental Health Center. All participants received psychiatric clinical evaluation, and the study focused on adolescents meeting DSM-5 diagnostic criteria for anxiety disorders or depressive disorders. The final multi-omics modeling cohort contained 107 participants, including 53 non-NSSI adolescents and 54 adolescents with NSSI.

NSSI was defined as deliberate, direct self-injury to body tissue without explicit suicidal intent and not socially sanctioned. Participants with two or more NSSI episodes within the previous six months were assigned to the NSSI group based on clinical interview and behavioral records. The non-NSSI comparison group consisted of adolescents with anxiety or depressive disorders but without NSSI or suicidal behavior.

Inclusion criteria were age 9-15 years, DSM-5 anxiety or depressive disorder diagnosis, availability of analyzable multi-omics, clinical, scale or MRI data, and written informed consent from participants and legal guardians. Exclusion criteria included DSM-5 psychotic disorder, current manic episode, autism spectrum disorder, intellectual disability, tic disorder, substance abuse, severe organic brain disease, severe physical illness, clear suicidal ideation, suicide attempt or suicidal behavior within the previous month, or severe missingness in key identifiers, clinical group labels or omics data. Clinical assessments included NSSI status and exploratory symptom dimensions, including impulsivity, somatic symptoms and OCD-related scores where available. MRI was available in a subset and was analyzed separately as exploratory brain-end anchoring. The study was approved by the Ethics Committee of Shanghai Mental Health Center (approval number 2022-74C3), and all personal information was de-identified before analysis.

Exploratory scale-derived symptom dimensions were calculated from Chinese versions of standardized self-report scales. Reverse-coded items were recoded before scoring so that higher values consistently indicated greater symptom burden or risk. Impulsivity was represented by the item-level mean score across Barratt Impulsiveness Scale (BIS) items. The somatic-symptom-related dimension was calculated by first deriving the item-level mean score within each of three scales, namely the Childhood Anxiety Sensitivity Index (CASI), the Screen for Child Anxiety Related Emotional Disorders (SCARED), and the Adolescent Sleep Scale, and then averaging these three scale-level mean scores. Among participants who completed a given scale, no within-scale item-level missing values were present; differences in sample size across symptom-association analyses, such as n = 67 for impulsivity and n = 80 for somatic symptoms, reflected differences in scale completion across participants rather than internal missingness within completed scales.

### 4.2 Multi-omics data acquisition and preprocessing

Genome-derived features included a polygenic risk score (PRS) and rare variant burden. Whole-genome sequencing data were not entered into the classifier as raw high-dimensional variants. Instead, common-variant genetic background was summarized using a two-stage MTAG-enhanced GWAS signal plus PRS-CSx cross-population modeling strategy. NSSI GWAS summary statistics were enhanced by MTAG using related self-harm phenotypes, including self-harm ideation, self-harm behavior and suicide attempt; the broader suicide-attempt genetic context was informed by UK Biobank-based studies [23, 36]. Following the PRS-CSx framework, two discovery summary-statistic files were supplied with population-specific LD reference panels to estimate ancestry-specific posterior SNP effect sizes [25]. The European-ancestry input was the MTAG-enhanced NSSI summary statistics (6,810,968 SNPs; N = 133,620; POP = EUR), and the East Asian input was the CONVERGE major depressive disorder summary statistics (4,879,285 SNPs; N = 10,640; POP = EAS) [57]. The PRS-CSx LD reference directory included 1000 Genomes EUR and EAS LD blocks and the multi-population HapMap3 SNP information file. PRS-CSx was run with phi = 1e-2, n_iter = 2000, n_burnin = 1000 and thin = 5. This genome-aware strategy was motivated by recent NSSI and suicide-genetics findings [22, 24].

For participant i, the ancestry-specific PRS was represented as a weighted sum of allele dosages:

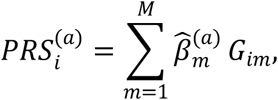

Where *a* denotes ancestry-specific effect estimates, *G_im_* is the allele dosage of variant *m* in participant *i*, and 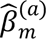 is the posterior SNP effect estimated by PRS-CSx. After harmonization and scoring, EUR and EAS profile scores were generated separately. The final PRS feature used in the model was an integrated weighted score:

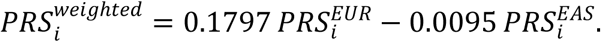

The scoring stage yielded EUR and EAS profile scores for 114 samples before intersection with the full multi-omics modeling matrix. Rare variant burden was derived from whole-genome sequencing by filtering rare exonic variants with allele frequency < 0.001 or allele count <= 5, annotating variant consequence with SnpEff/SnpSift and dbNSFP [37, 38], and prioritizing predicted damaging variants. High-priority variants included protein-truncating variants and damaging missense variants, defined using CADD > 20 [39], PolyPhen-2 damaging annotation [40] and MPC score > 3 where available [41]. Moderate-priority missense variants were defined by MPC score 2-3 [41]. Variants were further restricted to loss-of-function-intolerant genes (pLI > 0.9) [42] and genes related to brain function or psychiatric disorders. Individual-level constrained rare burden was then summarized as the rare burden feature.

Rare burden was summarized as a count-based constrained burden:

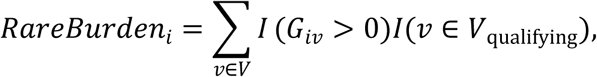

where denotes rare variants passing the frequency, functional-damage, pLI and brain-or psychiatric-gene filters.

Metabolomic profiling was generated from two main experimental batches. Raw mass-spectrometry outputs were processed for peak extraction, metabolite detection and quantitative feature generation, and annotated metabolites were mapped to HMDB and KEGG where available [43, 44]. Level 1 and level 2 annotated metabolites were retained for downstream analysis. Quality-control samples were used to assess reproducibility through principal component analysis and coefficient-of-variation summaries. Metabolomic data were log-transformed where applicable, normalized by probabilistic quotient normalization, corrected for signal drift using QC-based robust LOESS signal correction, and scaled before modeling [45–47]. No metabolomic missing values were identified after preprocessing.

Inflammatory proteins were measured using the Olink inflammation panel, containing 96 inflammation-related proteins covering cytokines, chemokines, immune-cell communication, inflammatory regulation and repair-related signals [48]. Inflammatory features were checked for completeness and scaled before modeling. Clinical blood indicators were obtained from routine clinical tests and covered neuroendocrine, immune, glucose-lipid metabolism and other baseline physiological indicators. Blood indicators overlapping with metabolomic or inflammatory measurements were removed to reduce redundant biological information. Missing blood values were median-imputed. Interval-type records were converted numerically: values coded as “max” were replaced with 1.1 times the maximum observed value of the corresponding indicator, and values coded as “min” were replaced with 0.9 times the minimum observed value. Continuous features were z-score standardized. Proximity-extension proteomic studies provided methodological precedent for this type of inflammatory-protein profiling [49, 50].

Samples were aligned by unified sample identifiers before modeling. Metabolomic, inflammatory, blood, PRS, rare burden and label tables were intersected to generate the final modeling matrix. Preprocessing steps that could learn from the data distribution or labels, including imputation, scaling, feature screening and model fitting, were performed within the training data during repeated model evaluation to reduce information leakage.

Continuous features were standardized within the training data as:

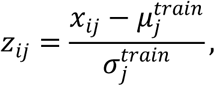

where *x_ij_* is the raw value of feature *j* in participant *i*, and 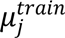 and 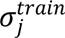 are the training-fold mean and standard deviation. The same training-fold parameters were then applied to validation or test samples.

### 4.3 Multi-omics expert models and stacking fusion

The classification pipeline used modality-specific expert models followed by a second-layer stacking logistic regression meta-model. Metabolomic and inflammatory features were each modeled using two complementary expert paths: random forest and ElasticNet logistic regression. Clinical blood baseline features were modeled using a random forest expert. PRS and rare burden were treated as genetic context variables and entered the second-layer model rather than being used as raw high-dimensional genomic inputs [51, 52].

Random forest experts used class-balanced training and 100 trees; the blood random forest used a maximum depth of 5. ElasticNet logistic regression used the SAGA solver, balanced class weights, five-fold internal cross-validation and candidate l1 ratios of 0.5 and 0.7. Voting-based feature selection was implemented within the training data for the feature-selection branches shown in Fig. 2a. The selector combined univariate group evidence from Mann-Whitney U tests, ElasticNet support and random forest importance support, and retained features supported by at least two evidence sources. The final meta-classifier was an L2-penalized logistic regression model with balanced class weights, C = 0.1, solver = ’lbfgs’, max_iter = 100, tol = 1e-4 and random_state = 42.

For the single-modality genomic benchmark, PRS and rare burden were used to train a logistic-regression classifier under the same repeated split setting.

For ElasticNet logistic regression, the fitted coefficients minimized the penalized negative log-likelihood:

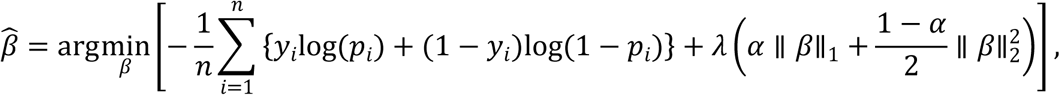

with

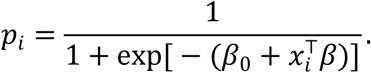

Here,*a* controls the balance between L1 and L2 penalties, and λcontrols the regularization strength.

For stacking, first-layer expert predictions were generated as out-of-fold probabilities using stratified five-fold cross-validation within the training data. In each inner fold, expert pipelines were cloned and refit on the corresponding inner-training subset; imputation, feature screening and model fitting were therefore performed within the inner-training data before predicting the inner-validation samples. These expert probability features were concatenated with scaled PRS and rare burden values. Genetic-by-omics interaction terms were constructed by multiplying each expert probability by each genetic context variable. The meta-classifier was trained on these out-of-fold expert probabilities, genetic context terms and interaction terms. Final base experts were then refit on the full training fold and applied to the held-out test data. Model performance was summarized over 20 repeated stratified random train-test splits using AUC, AUC variance, Brier score and related classification metrics. Each split used test_size = 0.20, yielding 22 held-out participants with balanced group representation (11 NSSI and 11 non-NSSI).

Let *^p_ik_* denote the out-of-fold predicted probability from expert model *k* for participant *i*, and let *g_il_* denote genetic context feature *l*. The stacking input vector was:

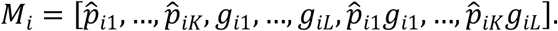

The second-layer model then estimated:

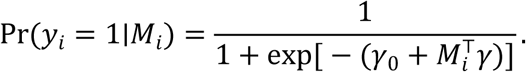

### 4.4 Repeated modeling and stable feature selection

Stable feature selection was performed through repeated modeling. The modeling procedure was repeated 20 times, and selected features were recorded across runs. A stable feature was defined as a feature selected in at least 80% of runs, corresponding to selection in 16 or more of 20 repeated runs. This procedure yielded 42 stable features for downstream interpretation.

For feature *j*, selection frequency was calculated as:

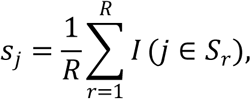

where *R* = 20 repeated runs and is the selected feature set in run r. A feature was defined as stable when:

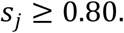

For each stable feature, univariate group comparisons between NSSI and non-NSSI participants were performed using two-sided Mann-Whitney U tests. Multiple testing was controlled using the Benjamini-Hochberg false discovery rate procedure [53]. Stable features were categorized by omics source. Meta-model contribution was summarized from the absolute coefficients of feature groups in the stacking layer, including omics expert probabilities, genetic context variables and genetic-by-omics interaction terms. For feature or group , the coefficient-based contribution was calculated as:

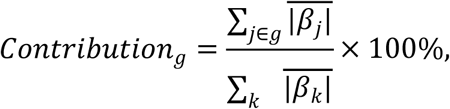

where 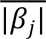 denotes the mean absolute logistic-regression coefficient across repeated modeling runs. These descriptive summaries were used to distinguish stable multivariate model features from conventional univariate biomarkers.

### 4.5 Group-specific network construction

Group-specific networks were constructed separately for the NSSI and non-NSSI groups using the 42 stable features; PRS_score and Rare_Burden were displayed as additional genetic-context nodes in Fig. 3a and were not counted toward the 42-feature total. Within each group, pairwise Spearman correlation coefficients were calculated across features. An undirected edge was added when the absolute Spearman correlation exceeded 0.30. Edge sign was retained to distinguish positive and negative associations, and absolute correlation magnitude was used as edge weight for topology summaries and visualization. These correlation networks were interpreted as feature-coordination networks rather than causal molecular interaction networks.

For features *u* and in group *g*, the edge rule was:

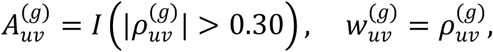

Where 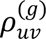 is the within-group Spearman correlation. Positive and negative edges retained the sign of 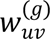, while 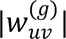 was used for weighted topology calculations.

Network visualization was used to compare global architecture, edge organization, module-level centrality reconfiguration, module transition and feature coordination across groups. Global summaries included edge turnover, largest connected component ratio, cross-omics edge proportion and top-hub overlap. Hub overlap was summarized using the Jaccard index of the top 10 betweenness hubs. The purpose of this analysis was to evaluate whether stable features were reorganized in the NSSI state rather than to infer causal molecular interactions [54].

Edge turnover was summarized as:

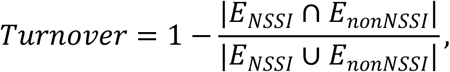

and top-hub overlap was summarized using:

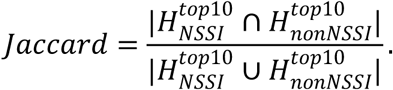

### 4.6 Module detection and network rewiring analysis

Community detection was performed on the group-specific networks using greedy modularity community detection weighted by absolute edge strength. A parallel Leiden-based community detection analysis yielded identical module organization, and the manuscript reports the greedy modularity partition to maintain a single consistent module nomenclature. Module assignment was compared between non-NSSI and NSSI networks. Module transition matrices and row-normalized flow ratios were used to quantify feature reassignment across groups.

Module-level centrality reconfiguration was summarized using betweenness centrality. For weighted betweenness, edge distance was defined as the inverse of absolute edge weight, 1/ max(|ρ|, 10^−9^), so that stronger correlations represented shorter network distances. For each feature, betweenness centrality was calculated in the group-specific networks, and between-group change was summarized as the absolute centrality shift. For a module , module-level reconfiguration was calculated as the sum of absolute betweenness shifts across module members, 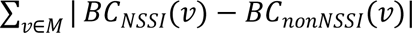 Modules with larger centrality reconfiguration values were interpreted as regions with stronger topological role shifts and were prioritized for downstream biological inspection together with module-transition evidence.

Betweenness centrality for node was defined as:

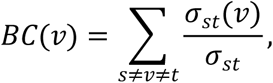

where σ_st_ is the number of shortest paths between nodes *s* and *t*, and σ*_st_*(*v*) is the number of those paths passing through *v*. Weighted shortest paths used:

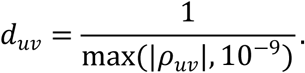

Module-level centrality reconfiguration was:

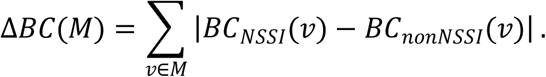

Edge-level rewiring analyses focused on biologically interpretable feature pairs within the reconfigured NSSI-M0-centered subnetwork. Within each group, pairwise associations were summarized using Spearman rho and corresponding P values. Group-specific coupling was tested using linear interaction models of the form *y*∼ *x* + group + *x*: group , and the interaction P value was used to summarize whether group status modified the feature-feature relationship. Regression panels used linear fits for visualization and interaction P values to describe group-specific coupling differences for selected ADA-, CXCL5-and NTF3-centered relationships.

### 4.7 MRI preprocessing and white-matter-normalized limbic readout extraction

MRI analyses were performed in the available MRI subset as exploratory brain-end phenotypic anchoring. After imaging quality control, sample matching and outlier removal, 19 participants were retained for the MRI analysis, including 6 non-NSSI and 13 NSSI participants. Raw MRI images were organized as NIfTI files and matched to clinical group labels and study identifiers. T1 images were bias-field corrected, skull-stripped, and registered to the MNI152 template before atlas-based ROI extraction. ROI extraction used the Harvard-Oxford subcortical atlas through Nilearn, and mean intensity values were extracted for left and right hippocampal and amygdala ROIs with NiftiLabelsMasker. Participant-specific white-matter masks were generated using FSL FAST segmentation, and the Harvard-Oxford atlas and participant-specific white-matter masks were brought into the same analysis space before extracting ROI and white-matter mean intensities. Each ROI readout was then expressed as a dimensionless white-matter-normalized ratio [55, 56]:

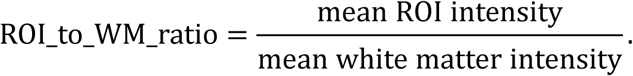

The displayed limbic analyses focused on Left Hippocampus_to_WM_ratio, Right Hippocampus_to_WM_ratio and Mean Hippocampus_to_WM_ratio. This normalization used white matter from the same image as an internal reference to reduce the influence of global brightness differences and scanner-related absolute intensity drift. These ratios were interpreted as relative signal-intensity features rather than physical-unit or quantitative MRI parameters.

Group comparisons were performed between NSSI and non-NSSI MRI subset participants using two-sided Mann-Whitney U tests. Peripheral support-related features, including D-serine, NTF3 and TNFSF12/TWEAK, were examined in relation to hippocampus-to-WM ratio readouts. Two-dimensional separation plots were used to visualize whether peripheral support-related features aligned with hippocampus-to-WM ratio gradients in the MRI subset. These MRI analyses were used for cross-scale phenotypic anchoring of the peripheral support-related signature.

### 4.8 Gut-related and stress-endocrine feature analyses

Gut-related and stress-endocrine analyses were performed as exploratory symptom-feature anchoring. Gut-related metabolites included stercobilin and trans-3-indoleacrylic acid, and steroid or endocrine-related features included cortisol, DHEA-S, aldosterone, 16alpha-OH-DHEA-S, and 5alpha-pregnan-3beta,20beta-diol 3-sulfate where available.

Interaction analyses examined whether relationships among steroid-related metabolites, gut-related metabolites and inflammatory features differed between groups. For interaction plots, linear models included the molecular feature, group and feature-by-group interaction term, and the interaction P value was used to summarize group-specific coupling. Symptom associations were assessed using Spearman correlations for impulsivity, somatic symptoms, OCD-related scores and other available scale dimensions.

A direction-aligned hormone burden score was calculated as a control-referenced, direction-aligned z-score average. Candidate stress-endocrine features included endocrine-related variables among the stable features. Each feature was standardized relative to the non-NSSI control mean and standard deviation, multiplied by a direction indicator, and averaged across included features. The direction indicator was defined as +1 for features higher in NSSI and -1 for features lower in NSSI.

Equivalently, for participant :

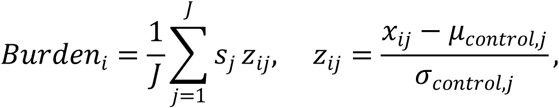

where *s_j_*=+ 1 if feature *j* was higher in NSSI and *s_j_*=− 1 if feature *j* was lower in NSSI.

### 4.9 Sentinel marker panel construction and case selection

The sentinel marker panel was constructed from model-prioritized features anchored in the NSSI-M0-centered network interpretation and the stress-endocrine context analysis. Within the NSSI-M0-centered module, markers with clear group separation were retained to represent the inflammatory, immune-communication and support and remodeling components of the network. To include a clinically accessible endocrine dimension, the strongest stress-endocrine signal was added as the cortisol/DHEA-S (C/D) ratio. Final markers were chosen to provide non-redundant functional coverage and profile-level clinical readability, because the panel was intended to summarize the full-model output rather than replace the full classifier.

To evaluate how much information was retained by the compact panel, a panel-only L2-regularized logistic regression model was trained using the selected sentinel markers. The model used balanced class weights, C = 0.1, the lbfgs solver, maximum iterations of 100, tolerance of 1e-4 and random_state = 42, matching the relevant meta-classifier settings. The panel-only model was evaluated across the same 20 repeated splits as the full model. AUC values from the panel-only model were compared with the corresponding full-model AUC values to benchmark the simplified panel against distributed multi-omics integration.

Representative cases were selected to illustrate the interpretive role of the panel. The displayed cases included a reference non-NSSI participant with a low full-model percentile, a reference NSSI participant with a higher full-model percentile, a panel-discordant NSSI participant correctly identified by the full model and an MRI-anchored NSSI participant with available hippocampus-to-WM ratio readout. For each case, full-model percentile and sentinel-marker percentiles were displayed; PRS or hippocampus-to-WM ratio percentiles were added where relevant. Case examples were interpreted as profile visualizations rather than independent validation of individual-level decision rules.

### 4.10 Statistical analysis

Unless otherwise specified, group comparisons used two-sided Mann-Whitney U tests. Correlations used Spearman rank correlation. Multiple testing was controlled using the Benjamini-Hochberg false discovery rate procedure where a family of related tests was performed [53]. Regression and interaction analyses used ordinary least-squares linear models for exploratory coupling analyses and visualization. Effect sizes, confidence intervals, P values and FDR-adjusted values were reported where available.

For Mann-Whitney U tests, observations from the two groups were ranked jointly. For the NSSI group with sample size _1_, the U statistic was:

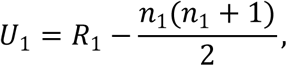

where *R*_1_ is the sum of ranks in the NSSI group. Two-sided P values were reported.

For Benjamini-Hochberg correction, P values were sorted as _(1)_ ≤ ⋯ ≤ _()_ , and adjusted values were summarized as:

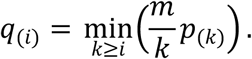

Machine learning analyses were implemented in Python using scikit-learn. Statistical analyses used SciPy and statsmodels, network analyses used NetworkX [54], and MRI ROI extraction used Nilearn with FSL FAST used for white-matter segmentation [55, 56]. Random seed 42 was used to initialize seeded model components and the repeated split generator; the evaluation still consisted of 20 distinct stratified train-test splits. Model performance was summarized using AUC and related metrics across repeated runs. Network-level, module-level, MRI and sentinel-panel case analyses were treated as exploratory and interpreted as evidence of group-specific feature coordination and state-profile readability rather than causal biological interactions.

## Data availability

The individual-level clinical, psychiatric, genomic, metabolomic, inflammatory-protein and MRI data generated in this study are not publicly available. The study involved minors with mental disorders, and the datasets contain sensitive health, behavioral, genetic and neuroimaging information. Because combining these high-dimensional data may create a risk of participant re-identification even after de-identification, unrestricted public disclosure would be inconsistent with the privacy protections, informed-consent conditions and ethical requirements governing this study. Requests for access may be directed to the corresponding authors and will be considered on a case-by-case basis in accordance with the original ethical approval, participant consent and applicable institutional policies.

## CRediT Author statement

**Fangyi Zhao**: Conceptualization, Methodology, Software, Formal analysis, Investigation, Data curation, Validation, Visualization, Project administration, Writing – original draft, Writing – review & editing. **Yihang Bao**: Conceptualization, Methodology, Supervision, Project administration, Writing – review & editing. **Wenjing Liu**: Investigation, Data curation. **Tianqi Liu**: Methodology, Software. **Weidi Wang**: Conceptualization, Methodology, Writing – review & editing. **Zhen Liu**: Conceptualization, Methodology, Writing – review & editing. **Xiaoxia Lei**: Conceptualization, Methodology, Writing – review & editing. **Xifeng Xia**: Conceptualization, Methodology, Writing – review & editing. **Wenhong Cheng**: Conceptualization, Data curation, Supervision, Project administration, Resources. **Guan Ning Lin**: Writing – review & editing, Validation, Supervision, Project administration, Investigation, Resources.

## Data Availability

https://opengwas.io/

https://figshare.com/articles/dataset/GWAS_summary_statistics_on_Major_Depressive_Disorder/3840696?file=10374393

