## Supplementary Materials for "Multi-Omics Modeling Reveals Peripheral Signatures of Non-Suicidal Self-Injury in Adolescents"

#### Supplementary Figures

**Supplementary Figure 1.** Model-performance summaries.
**Supplementary Figure 2.** Stable biomarker group differences.
**Supplementary Figure 3.** Stable-feature network comparison.
**Supplementary Figure 4.** Focused hub views.
**Supplementary Figure 5.** NSSI module composition and rewiring.
**Supplementary Figure 6.** Clinical and symptom comparison.
**Supplementary Figure 7.** Sentinel markers across model strata.
**Supplementary Figure 8.** Panel-only and full-model probabilities.
**Supplementary Figure 9.** Multi-omics correlation heatmaps.
**Supplementary Figure 10.** PRS and metabolic strata.
**Supplementary Figure 11.** Random Forest SHAP summaries.
**Supplementary Figure 12.** Hub-shifting mirror charts.

#### Supplementary Notes

**Supplementary Note 1.** How to read supplementary network figures.
**Supplementary Note 2.** Interpretation boundary for exploratory supplements.
**Supplementary Note 3.** Abbreviation and marker-display conventions.
**Supplementary Note 4.** Additional supplementary-figure methods

#### Supplementary Tables

**Supplementary Table 1**. Non-NSSI network-node module membership.
**Supplementary Table 2**. NSSI network-node module membership.

### Supplementary Figure

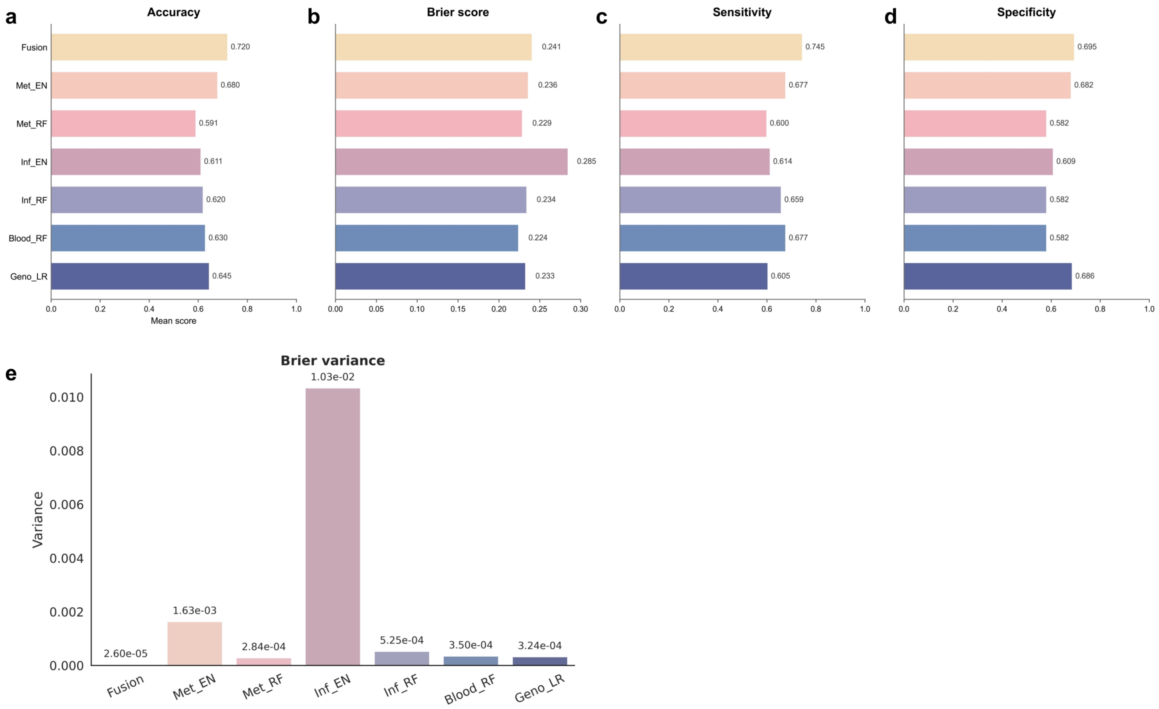


**Supplementary Figure 1.** Model-performance summaries. Accuracy, Brier score, sensitivity and specificity complement the main AUC comparison.

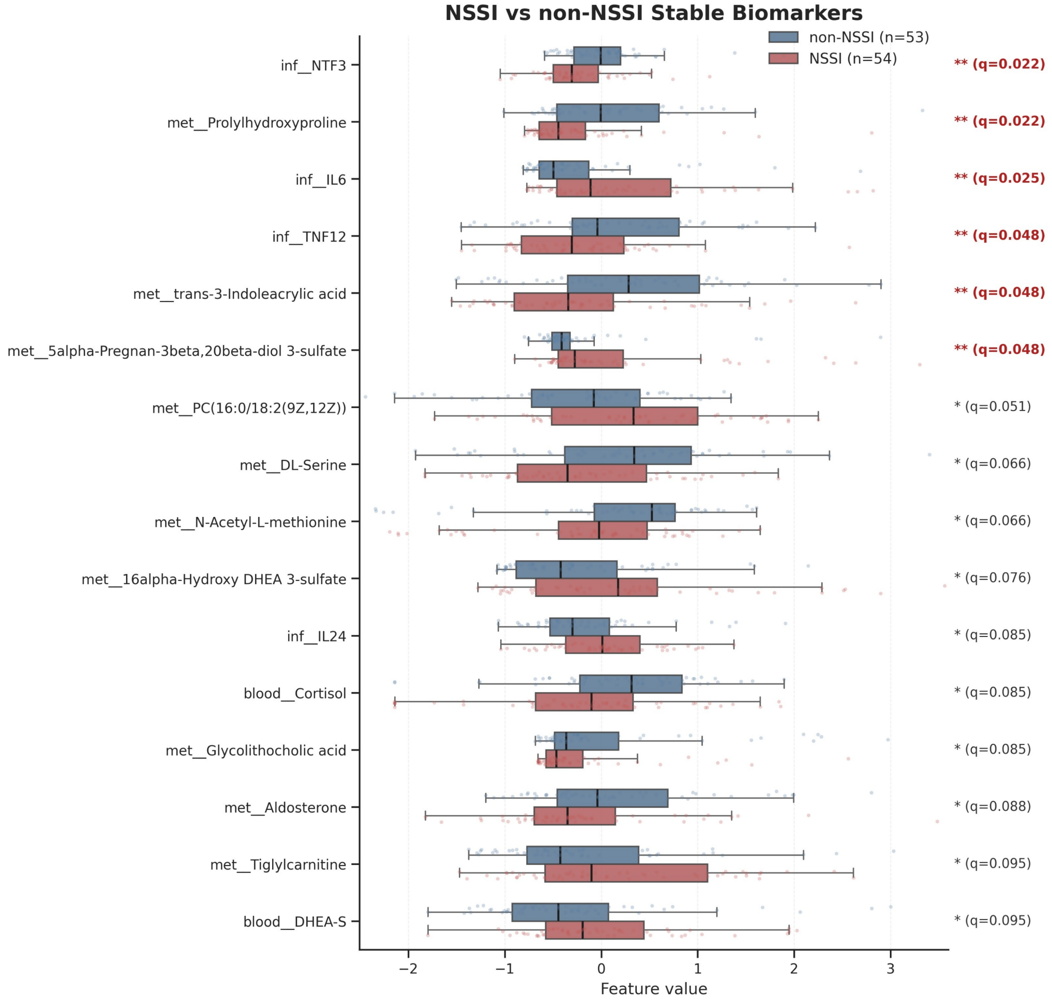


**Supplementary Figure 2.** Stable biomarker group differences. Box-and-jitter plots show group distributions for model-prioritized stable features.

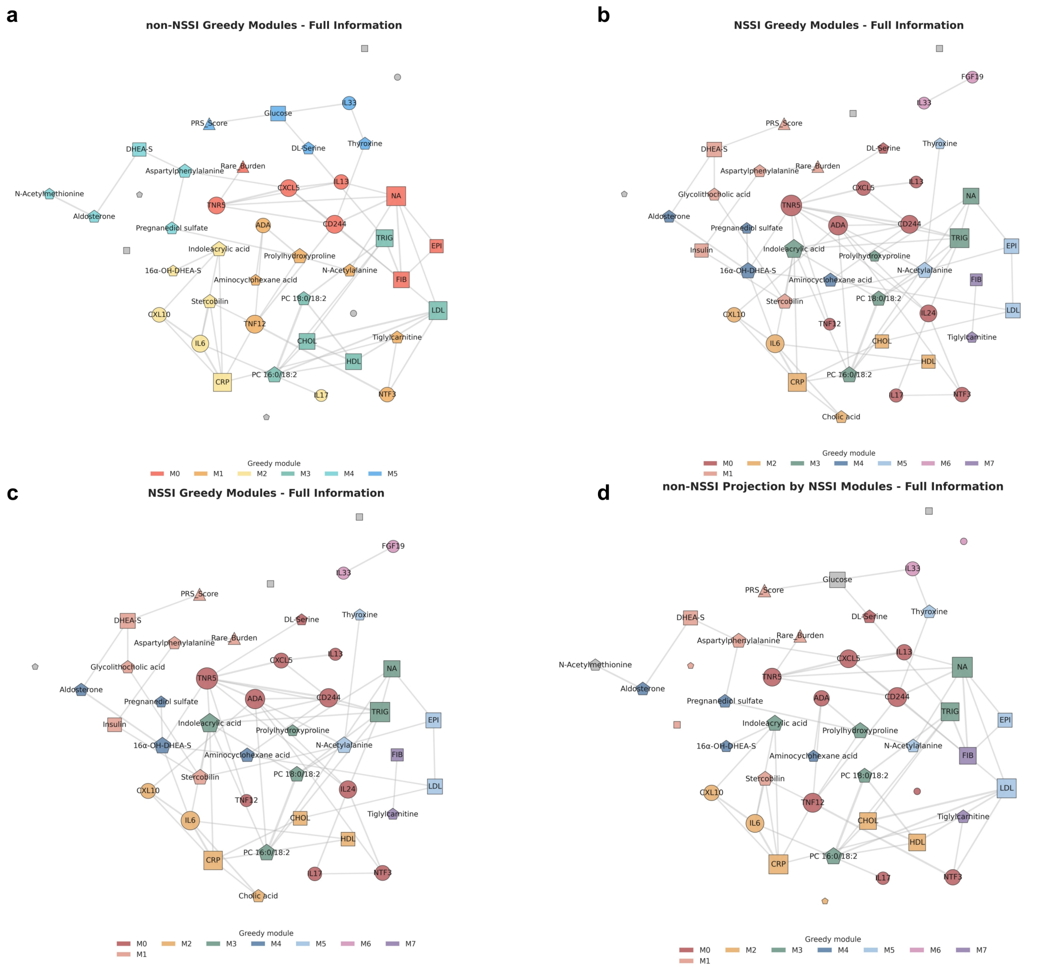


**Supplementary Figure 3.** Stable-feature network comparison. Full network views compare non-NSSI and NSSI module organization among stable model-prioritized features.

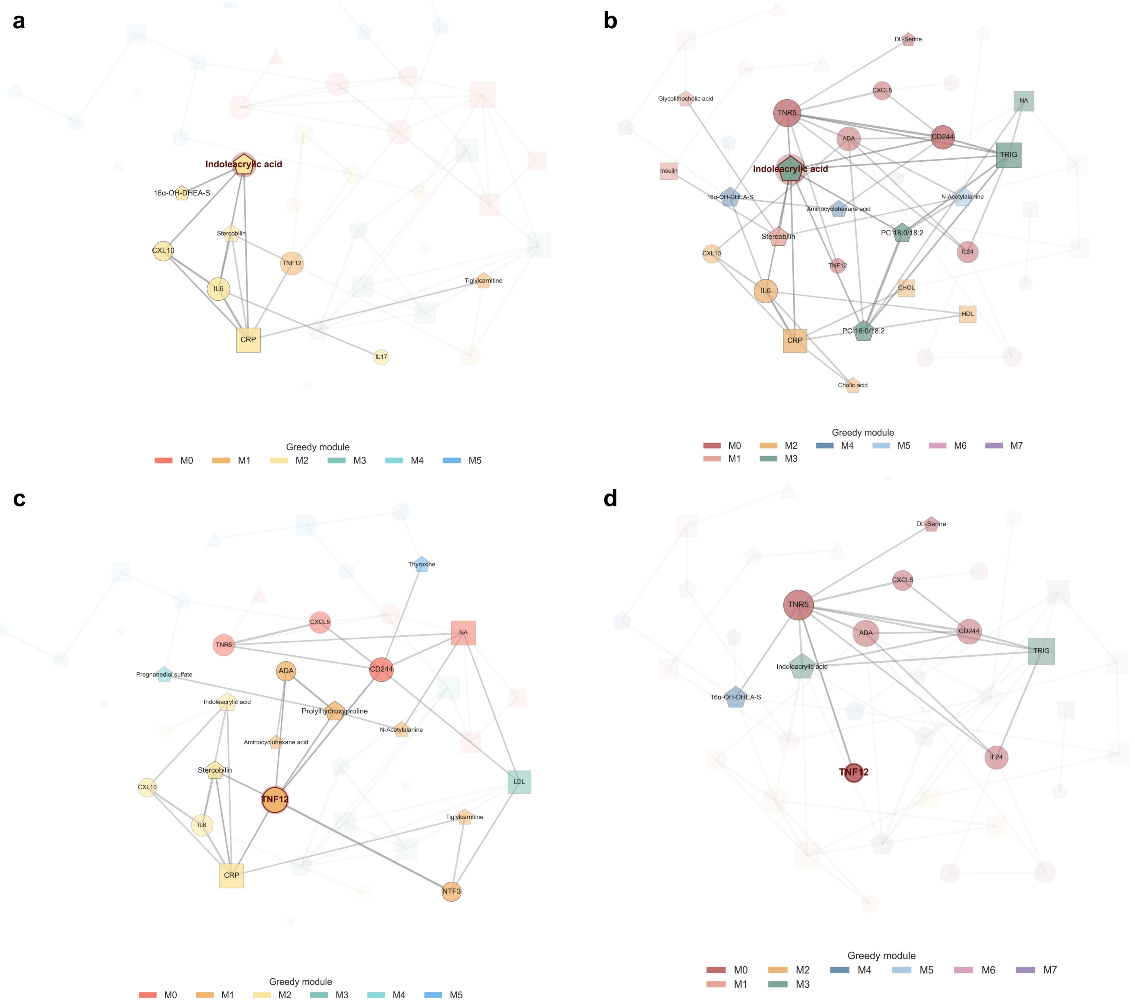


**Supplementary Figure 4.** Focused hub views. Local views show neighborhood structure around indoleacrylic acid and TNFSF12/TWEAK in the two group-specific networks.

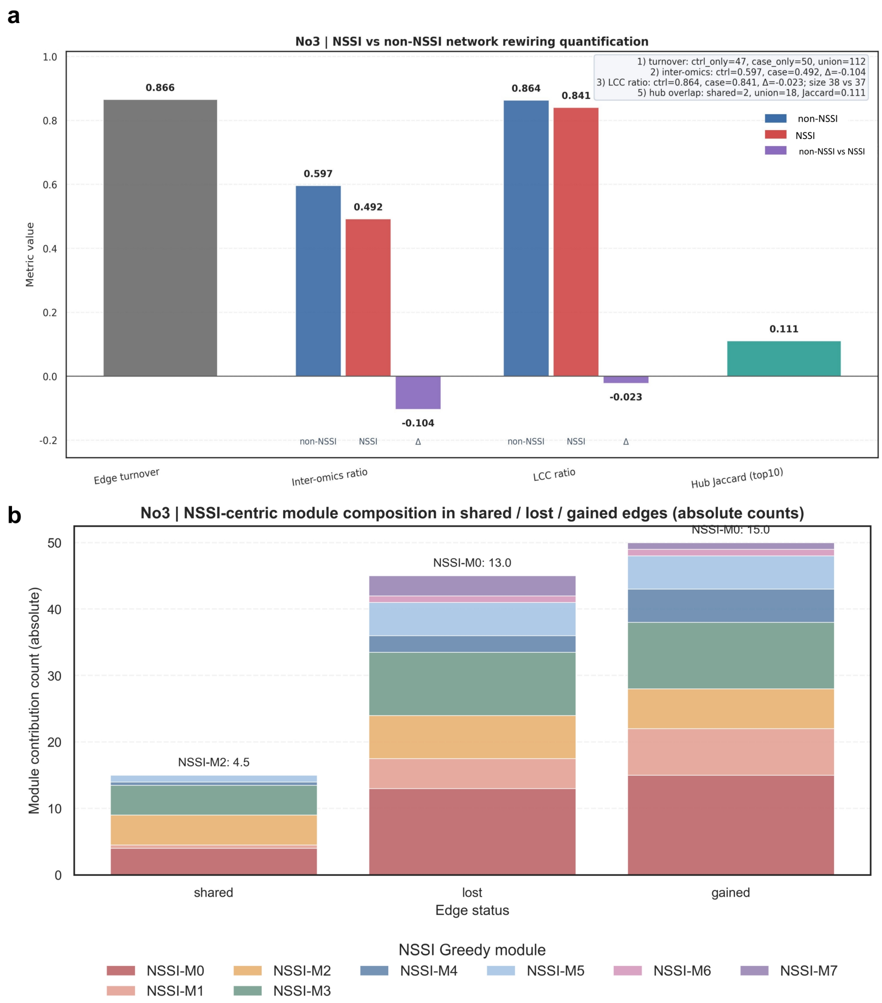


**Supplementary Figure 5.** NSSI module composition and rewiring. Panels summarize NSSI-centered module composition and network-level rewiring statistics.

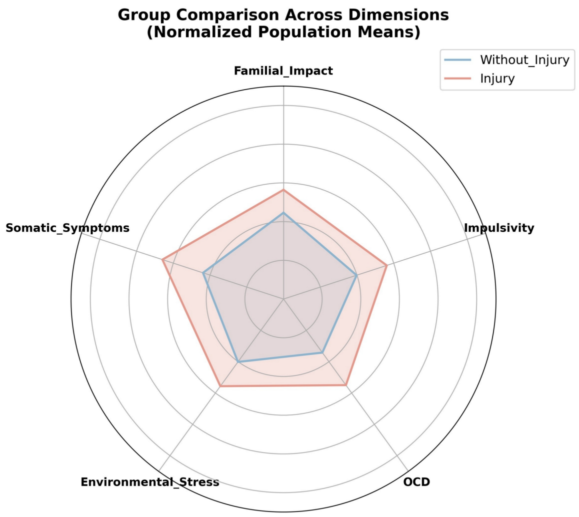


**Supplementary Figure 6.** Clinical and symptom comparison. Radar plots provide clinical-context summaries for symptom and clinical dimensions.

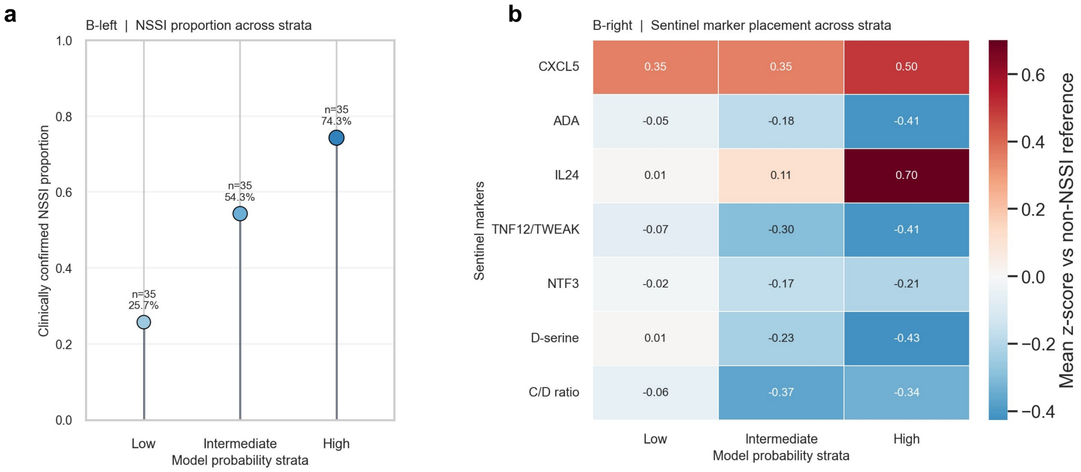


**Supplementary Figure 7.** Sentinel markers across model strata. Panels show NSSI proportion and sentinel-marker z-score profiles across full-model probability strata.

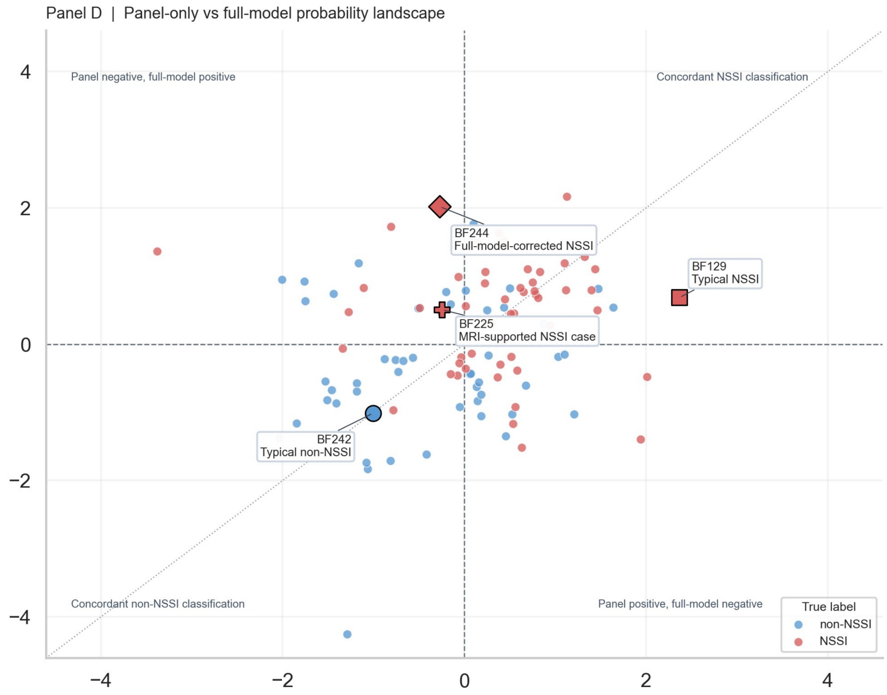


**Supplementary Figure 8.** Panel-only and full-model probabilities. Scatter plot compares the compact sentinel-panel model with the full genome-aware multi-omics model.

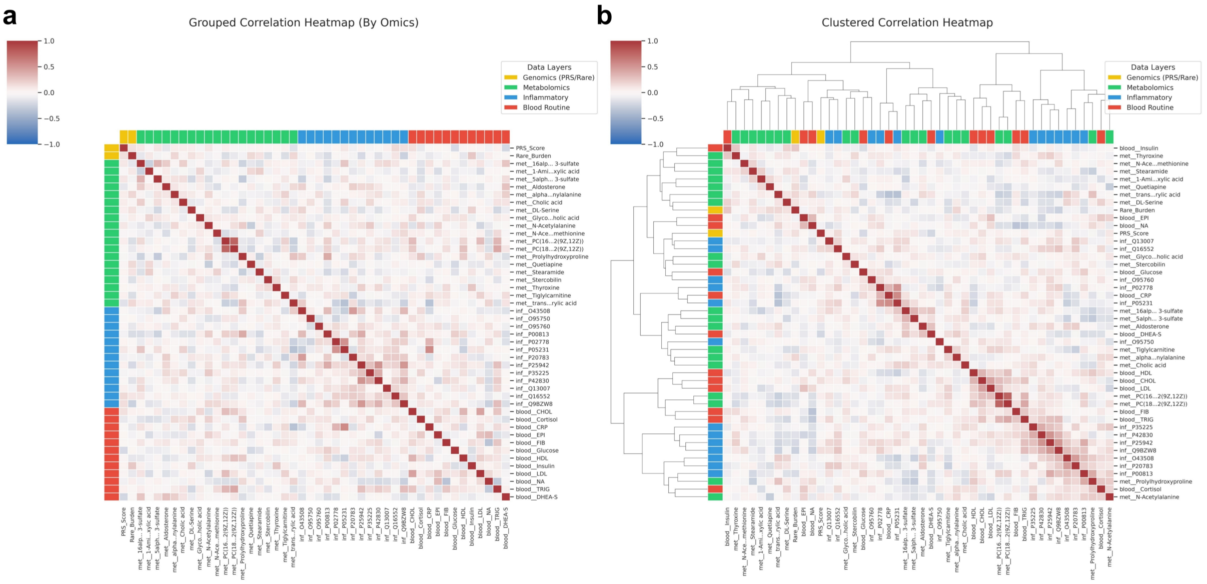


**Supplementary Figure 9.** Multi-omics correlation heatmaps. Grouped and clustered Spearman heatmaps show relationships among selected stable features.

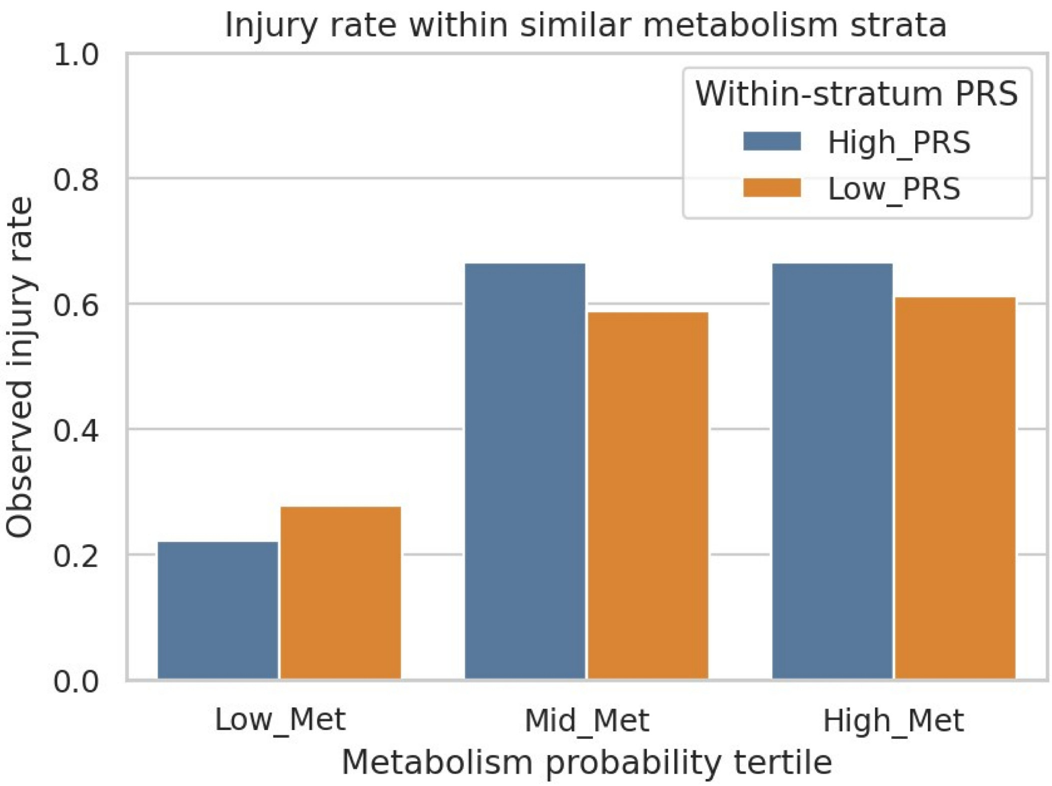


**Supplementary Figure 10.** PRS and metabolic strata. Observed injury/NSSI rates are shown after stratifying participants by metabolomics-derived probability and PRS group.

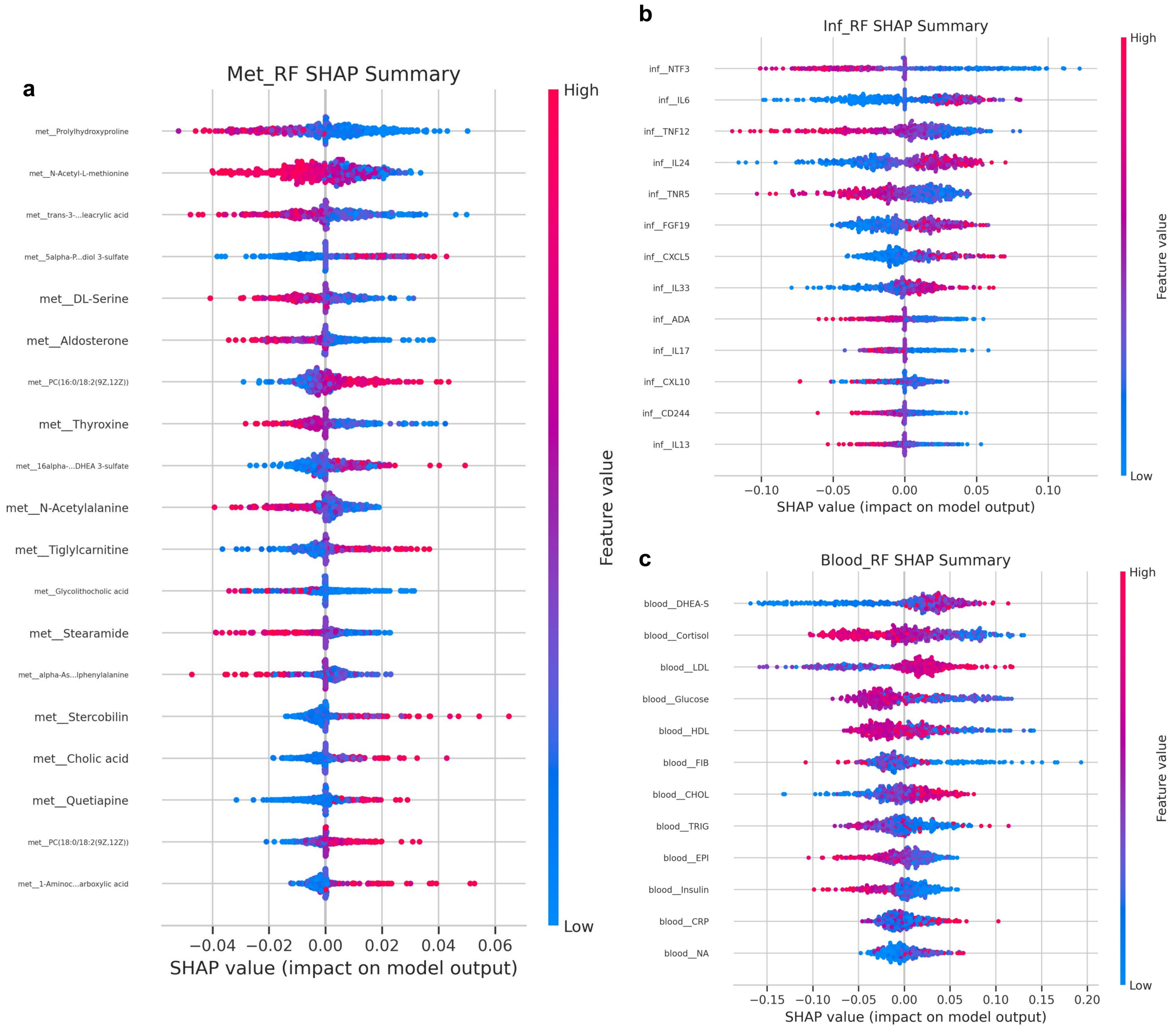


**Supplementary Figure 11.** Random Forest SHAP summaries. Seed-aggregated SHAP plots summarize feature contributions for metabolomic, inflammatory and blood expert models.

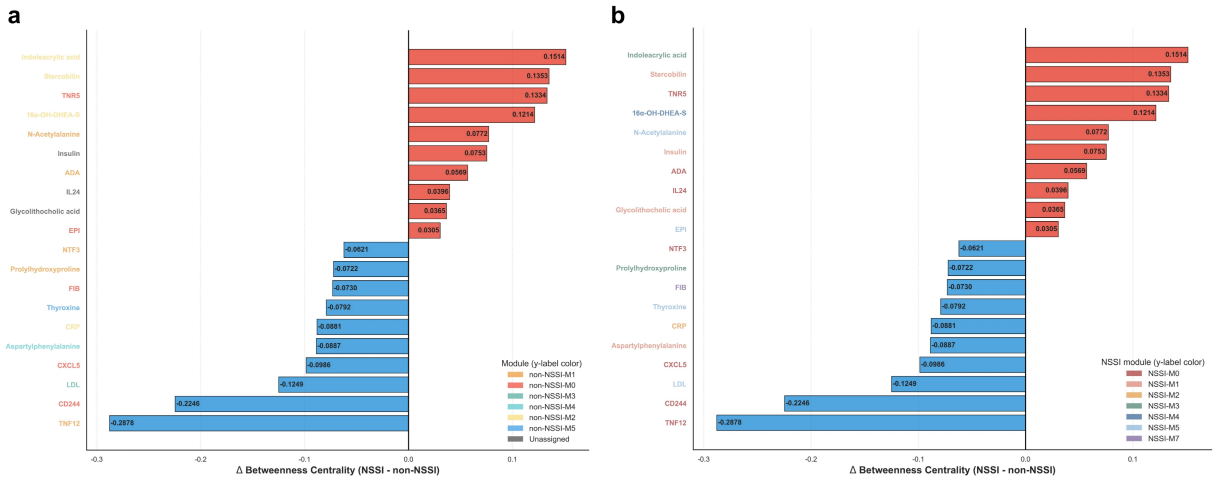


**Supplementary Figure 12.** Hub-shifting mirror charts. Delta betweenness centrality highlights features with increased or reduced bridge-like roles in the NSSI network.

### Supplementary Note

#### Supplementary Note 1. How to read supplementary network figures

The supplementary network figures are intended to expose the full graph context behind the simplified main-text panels. Module colors indicate community assignments within the displayed group-specific network. Projection panels should be read as cross-state maps of module labels rather than as causal transition diagrams.

#### Supplementary Note 2. Interpretation boundary for exploratory supplements

The supplementary figures extend model interpretation by showing additional diagnostics, local hub views and clinical-context summaries. These analyses support transparency and hypothesis generation, but they should not be interpreted as independent clinical validation or as evidence for causal molecular pathways.

#### Supplementary Note 3. Abbreviation and marker-display conventions

TNF12 denotes TNFSF12/TWEAK and TNR5 denotes TNFRSF5/CD40 where space is limited in network graphics. NSSI and non-NSSI colors follow the figure-specific legends. Feature values in distribution panels are displayed on the normalized analysis scale, whereas model-probability panels use classifier-derived probability or logit-scale summaries as indicated in the figure.

#### Supplementary Note 4. Additional supplementary-figure methods

Additional visualizations were generated for supplementary analyses not detailed in the main Methods. Multi-omics heatmaps used participant-aligned stable features and Spearman correlations. PRS-metabolism plots stratified metabolomics-derived out-of-fold probabilities into tertiles before comparing PRS groups. Random Forest SHAP summaries aggregated seeded expert-model explanations, and hub-shifting mirror charts displayed group differences in betweenness centrality.

### Supplementary Table

Supplementary Table 1. Non-NSSI network-node module membership.

| Module | Features | Feature list |
|---|---|---|
| M0 | 8 | CD244, CXCL5, EPI, FIB, IL13, NA, Rare_Burden, TNR5 |
| M1 | 7 | ADA, Aminocyclohexane acid, N-Acetylalanine, NTF3, Prolylhydroxyproline, Tiglylcarnitine, TNF12 |
| M2 | 7 | 16alpha-OH-DHEA-S, CRP, CXCL10, IL17, IL6, Indoleacrylic acid, Stercobilin |

| | | |
|---|---|---|
| M3 | 6 | CHOL, HDL, LDL, PC 16:0/18:2, PC 18:0/18:2, TRIG |
| M4 | 5 | Aldosterone, Aspartylphenylalanine, DHEA-S, N-Acetylmethionine, Pregnanediol sulfate |
| M5 | 5 | D-serine, Glucose, IL33, PRS_score, Thyroxine |
| Unassigned | 6 | Glycolithocholic acid, Cholic acid, IL24, FGF19, Insulin, Cortisol |

Supplementary Table 2. NSSI network-node module membership.

| **Module** | **Features** | **Feature list** |
|---|---|---|
| M0 | 10 | ADA, CD244, CXCL5, D-serine, IL13, IL17, IL24, NTF3, TNF12, TNR5 |
| M1 | 7 | Aspartylphenylalanine, DHEA-S, Glycolithocholic acid, Insulin, PRS_score, Rare_Burden, Stercobilin |
| M2 | 6 | CHOL, Cholic acid, CRP, CXCL10, HDL, IL6 |
| M3 | 6 | Indoleacrylic acid, NA, PC 16:0/18:2, PC 18:0/18:2, Prolylhydroxyproline, TRIG |
| M4 | 4 | 16alpha-OH-DHEA-S, Aldosterone, Aminocyclohexane acid, Pregnanediol sulfate |
| M5 | 4 | EPI, LDL, N-Acetylalanine, Thyroxine |
| M6 | 2 | FGF19, IL33 |
| M7 | 2 | FIB, Tiglylcarnitine |
| Unassigned | 3 | N-Acetylmethionine, Glucose, Cortisol |

Module membership follows Fig. 3a. The Unassigned rows list visible network nodes not assigned to a greedy community in that group. The network display includes PRS_score and Rare_Burden as genetic-context nodes; excluding these two genetic-context nodes leaves the 42 stable feature total used in the main text.
